# Distinct immune signatures are a potent tool in the clinical management of cytokine-related syndrome during immune checkpoint therapy

**DOI:** 10.1101/2024.07.12.24310333

**Authors:** Douglas Daoudlarian, Amandine Segot, Sofiya Latifyan, Robin Bartolini, Victor Joo, Nuria Mederos, Hasna Bouchaab, Rita Demicheli, Karim Abdelhamid, Nabila Ferahta, Jacqueline Doms, Grégoire Stalder, Alessandra Noto, Lucrezia Mencarelli, Valérie Mosimann, Dominik Berthold, Athina Stravodimou, Claudio Sartori, Keyvan Shabafrouz, John A Thompson, Yinghong Wang, Solange Peters, Giuseppe Pantaleo, Michel Obeid

## Abstract

Immune-related cytokine release syndrome (irCRS) frequently occurs during immune checkpoint inhibitor (ICI) therapy. In the present study, we have attempted to identify biomarkers in oncology patients experiencing irCRS-like symptoms (n=35), including 9 patients with hemophagocytic lymphohistiocytosis (irHLH)-like manifestations (8 classified as Grade (G) 4 irCRS and 1 as G3 irCRS) and 8 with sepsis, differentiating between irCRS, irHLH and sepsis. Patients grouped in three clusters based on distinct cytokine profiles and survival outcomes. We identified 24 biomarkers that significantly discriminated between irHLH and irCRS G3 (P < 0.0455 to < 0.0027). Notably, HGF and ferritin demonstrated superior predictive values over the traditional HScore, with a positive predictive value (PPV) and negative predictive value (NPV) of 100%. Furthermore, CXCL9 not only distinguished between irHLH and irCRS G3, but was also a predictor of treatment intensification with tocilizumab (TCZ) with a PPV of 90% and a NPV of 100%. Other parameters, such as leukocyte count, neutrophils, ferritin, IL-6, IL-7, EGF, fibrinogen, and GM-CSF, were effective in discriminating sepsis from high-grade irCRS with a PPV of 75-80% and an NPV of 100%. In comparison to sepsis, the frequencies of CXCR5+ or CCR4+ CD8 memory, CD38+ ITM monocytes, and CD62L+ neutrophils were observed to be higher in high-Grade irCRS. Of note, TCZ treatment led to complete resolution of clinical symptoms in 12 patients with high-grade irCRS refractory to corticosteroids (CS). These findings demonstrate the power of unique immunologic biomarkers in determining the severity of irCRS, in predicting survival, and distinguishing between high-grade irCRS, irHLH and sepsis. Therefore, these distinct unique signatures are instrumental for the optimal development of personalized clinical and therapeutic management in patients experiencing irCRS patient.

## Main

Immune Checkpoint Inhibitors (ICIs) have reshaped the anticancer landscape, showing remarkable efficacy across a spectrum of malignancies (*1*). However, ICIs are frequently associated with immune-related adverse events (irAEs) (*2*), which encompass a range of immune-mediated toxicities that raise substantial clinical challenges (*1, 3*). Among these, immune-related cytokine release syndrome (irCRS) is of particular concern due to its potential severity and life-threatening manifestations(*4*). irCRS is characterized by an acute, systemic inflammatory response, primarily mediated by an extensive release of cytokines. Historically, irCRS was associated with monoclonal antibodies (mAbs) such as rituximab, brentuximab and obinutuzumab and more recently with T-cell-engaging therapies (e.g., bispecific T-cell-engaging (BiTE) single-chain antibodies) and chimeric antigen receptor (CAR) T cells therapies. irCRS is now increasingly observed in the context of ICI therapy (*4–12*). Severe irCRS can lead to multi-organ dysfunction and, in extreme cases, can be fatal (*13*).

The clinical features and laboratory alterations of irCRS itself are not specific and may overlap or coincide with infectious diseases (*14*). In severe cases, irCRS may present with sepsis-like clinical signs and laboratory changes mimicking sepsis, macrophage activation syndrome or immune-related hemophagocytic lymphohistiocytosis (irHLH) (*15–17*). Importantly, a subgroup of patients treated with CAR-T cells (*18*) or BITE (*19*) can develop HLH-like features as a severe variant of CRS. In addition, CAR T-cell associated HLH (carHLH) identified as a CRS variant presented cytokine profiles and clinical manifestations like secondary HLH/macrophage activation syndrome (MAS) (*18, 20*). HLH can present with a wide spectrum of manifestations ranging from isolated biological abnormalities to a severe multi-organ clinical syndrome. Distinguishing between irCRS, especially in high grade (*15*), and sepsis (*21*) in patients receiving ICI therapy is crucial for effective clinical management.

An important unanswered dimension in the progression of irCRS is the extent to which the hyperinflammatory response evolves into or coexists with a compensatory anti-inflammatory response syndrome (irCARS) (*22–26*). This balance between pro-and anti-inflammatory forces is critical to understanding whether a pronounced anti-inflammatory phase could emerge in response to the inflammatory cascade seen in irCRS. The potential transition from an overwhelming inflammatory response to a compensatory anti-inflammatory phase could have profound implications for patients’ management and treatment outcomes. Furthermore, the possibility that high-grade irCRS may present with (HLH)-like manifestations adds an additional layer of complexity to the diagnosis and management of severe cases. Delineation of this continuum and its potential overlap with HLH-like presentations is essential to optimize therapeutic strategies and clinical outcomes.

A variety of grading systems for CRS have been proposed (*27–30*) with the objective of establishing standardized criteria. However, the current grading systems present several limitations and the diversity of criteria used are at the base of the inconsistency in evaluating of the safety profiles of different therapeutic agents, and the source of clinical misclassification. For instance, the performance of the HScore (*31*), traditionally used to estimate the individual risk of reactive HLH, may not be appropriate in the case of irHLH resulting from ICI-associated irAEs. Improving these grading systems and scores with precise immune measurements offer a promising avenue for refining the accuracy of severity grading and prognostic predictions, with the ultimate goal of rationalizing patient management and improving therapeutic outcomes.

In the present study, we investigated whether immunologic biomarkers can contribute to establishing solid criteria to apply to the clinical grading and classifications of ICI-associated irAES. For these purposes, comprehensive set of 115 biomarkers, including 50 cytokines, chemokines and growth factors and 44 cellular markers were analyzed. Our ultimate objective was to identify specific biomarkers that can discriminate between the different clinical manifestations of irCRS including irHLH and sepsis, and to predict severity, and patient survival. These findings are expected to facilitate the development of tailored therapeutic interventions.

### Clinical and demographic characteristics of the cohort

Of the 709 identified patients treated with ICI at the Lausanne University Hospital between 2020 and the end of 2023, 43 patients presented with clinical and biological presentation of systemic inflammatory response syndrome (SIRS) such as sepsis or irCRS according to ACCP/SCCM Consensus Conference Committee (*32, 33*) (**Supplementary Table 1**). Of these, 35 patients were included in the final analysis as described in the study flowchart (Supplementary Fig. 1). A subset of 28 patients were diagnosed with grades 1 to 4 of irCRS according to the Lee grading scale (*34*) and 7 patients were diagnosed with sepsis, microbiologically documented, including 2 cases of viral and 5 of bacterial sepsis. Extensive screening to exclude an infection as the cause of CRS was part of our local standard of care for irCRS (including HLH). Among the 28 patients with irCRS, 12 [43%] were classified as low-G irCRS (G1, G2) and 16 [57%] as high-G irCRS (G3, G4). Of note, 9 out of 28 (32%) met the criteria of reactive HLH according to the criteria (*31*). The use of five of the eight diagnostic criteria from HLH-2004 (*35*) has proven to be an effective tool for the diagnosis of irHLH. Adjustments to the HLH-2004 criteria, including setting hyperferritinemia cutoffs at 3000 μg/L and fever at 38.2°C, have increased sensitivity and specificity to 97.5% and 96.1%, respectively, as recently reported (*36*). All of our irHLH cases presented with hyperferritinemia greater than 3000 μg/L, even those with an HScore as low as 162, and showed conventional biological changes such as hepatitis, renal dysfunction, cytopenia, hypertriglyceridemia, hypofibrinogenemia, and hemophagocytosis in the bone marrow and spleen (although only three patients were biopsied). In addition, elevated levels of soluble CD25 (IL-2R) and key clinical features such as persistent fever and multi-organ dysfunction were observed in all cases.

Among the irHLH patients, 1 of the 9 patients was classified as irCRS G3 and 8 were classified as irCRS G4. None of the non-irHLH patients were classified as G4. Melanoma and lung cancer were the two most common types of cancer, with 17 out of 35 patients (49%) and 11 out of 35 patients (31%) respectively. Treatments modalities included anti-PDL-1 for 3 patients (9%), anti-PD-1 for 11 (32%), anti-PD-1 and anti-CTLA4 combination for 21 patients (60%) and chemo-immunotherapy for 9 patients (25.7%). The median follow-up after the first ICI administration was 29.5 months (95% CI: 13.68-44.44). Of the 28 patients with irCRS, 3 (11%) received corticosteroids (CS) alone, 13 (46%) received CS combined with tocilizumab (TCZ), and 12 (43%) did not receive any immunosuppressive treatment. Patients treated with anti-PD-1 and anti-CTLA4 combination therapy had a higher frequency of high-G (G3, G4) irCRS (75%, n=12/16) compared to those treated with anti-PD-1 monotherapy (25%, n=4/16). The median time to the onset of low-and high-G irCRS was 1.3 months (95% CI: 0.23-26.1) and 2.5 months (95% CI: 1.35-5.46), respectively.

### Comprehensive cytokine and biological profiles across different clinical grades of irCRS

To characterize the inflammatory profile of irCRS, we analyzed a large panel of classical and inflammatory biomarkers in patients with different clinical grades (G1 to G4). Of the 28 patients analyzed for irCRS, three were excluded from cytokine analysis due to ongoing immunosuppression at the time of samples collection. All other patients were screened for cytokines after referral to the Service of Immunology and Allergy prior to initiation of immunosuppression. Standard laboratory tests showed a significant association between irCRS severity and elevated levels of aspartate aminotransferase (AST), alanine aminotransferase (ALT), alkaline phosphatase (AP), gamma-GT (GGT), and ferritin, as well as decreased levels of fibrinogen (**Fig. 1a-1c**). Notably, coagulation abnormalities escalated with irCRS severity, characterized by elevated d-dimer levels and reductions in platelets, fibrinogen, prothrombin ratio, and activated partial thromboplastin time. The coagulopathy was particularly pronounced in grade 4 (G4) patients (**Fig. 1b-1c**).

**Figure 1.**
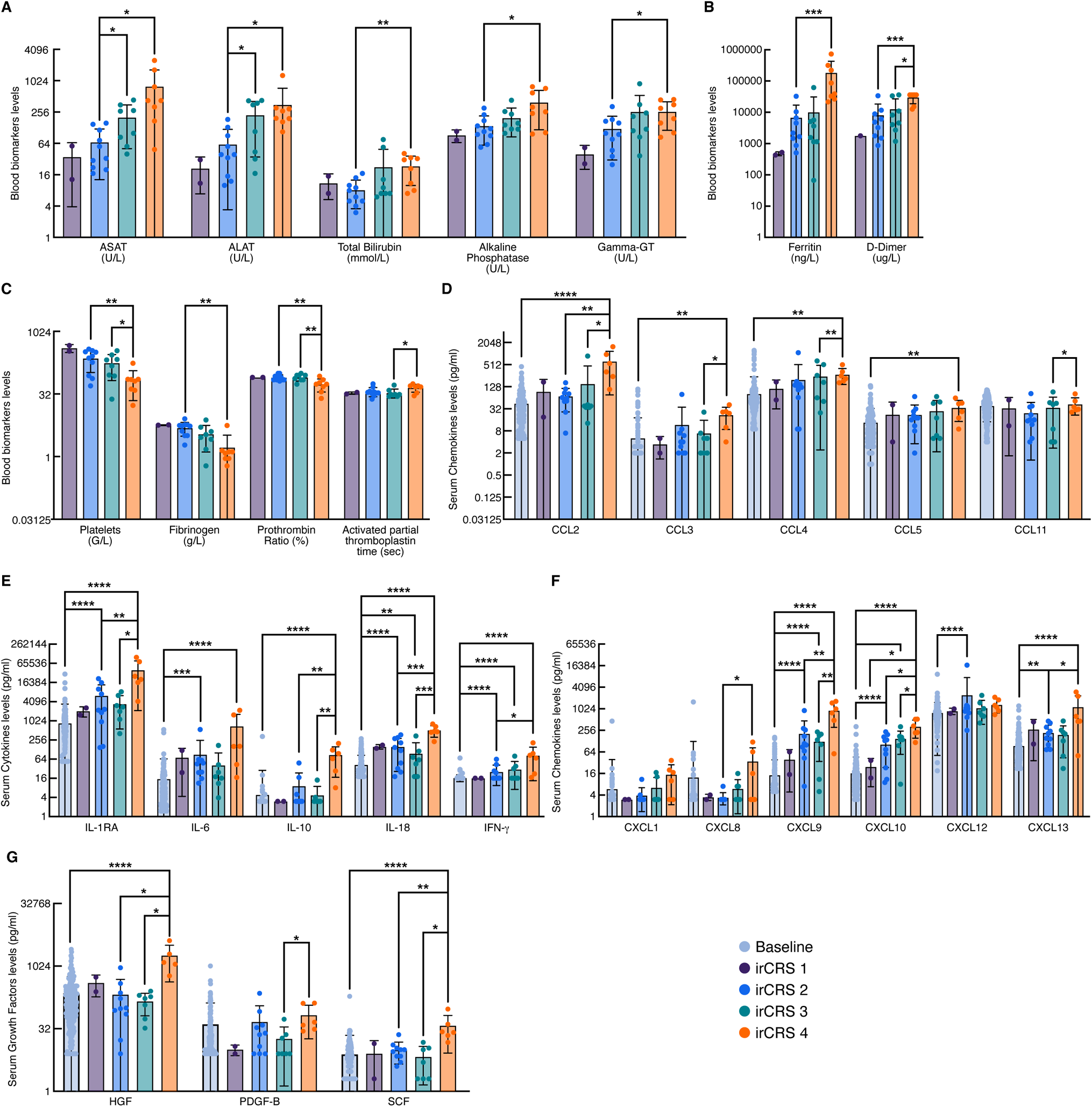
Comprehensive cytokine and biological profiles for different clinical grades of irCRS. (**a-c**), Evaluation of serum biomarkers in patients (n=28) at the time of irCRS compared to the pre-ICI serum levels in 194 cancer patients used as reference group. (d-g) Serum levels of chemokines, growth factors and cytokines in 25 patients with different clinical grades at irCRS diagnosis compared to the pre-ICI serum levels in 194 cancer patients used as reference group. Student T test tests were used to analyze the data for statistical significance between groups, irCRS G1 were not compared to other clusters due to the low number of patients (n=2). Plots represent values with individual data points, bar represent the mean and error bars represent standard deviation. The results showed a significant difference between the groups with a P-value of less than 0.001 (***P<0.001) and less than 0.0001 (****P<0.0001).

With regard to the analysis of the serum levels of a large panel (n=45) of cytokines/chemokines and growth factors, 14 were found to be increased as compared to baseline, i.e. prior to the initiation of ICI therapy and correlated with the irCRS grade severity. These included the chemokines CXCL9, CXCL10, CXCL13, CCL2, CCL3, CCL4, CCL5 and CCL11 (**Fig. 1d-1f**,), the growth factors SCF and HGF, (**Fig. 1g**), the inflammatory cytokines IL-6, IL-18 and IFN-γ, and the anti-inflammatory cytokines IL-1RA and IL-10. (Fig. 1**h**). Analysis of anti-inflammatory cytokines revealed an increase in IL-1RA in all irCRS grades with irCRS severity, and a significant increase in IL-10 levels in G4 (**Fig. 1e**).

By analyzing the correlation matrix of biomarkers across all irCRS patients, a complex network of positive and negative correlations was revealed. CRP showed a positive correlation with ferritin, IL-18, IL-6 and ALP, but a negative correlation with Hb. Inversely, Hb showed a negative correlation with IL-6 (**Supplementary Figure 2a**). Ferritin, a key marker of inflammation, correlated with a wide array of biomarkers, including PDGF-B, CCL2, HGF, SCF, CXCL13, CXCL9, IL-18, ALT, AST, CCL3, IL-1RA, CXCL10, CRP, CXCL1, IL-10, IFN-γ, and ALP (**Supplementary Figure 2**). Certain biomarker correlations were specific to patients with high-G and not observed in low-G such as IFNγ/IL-1RA, IL-10/HGF, IL-10/IL-1RA, IL-10/ferritin, d-dimer/ferritin, CRP/ferritin, HGF/ferritin and ALT/Hb **(Supplementary Figure 2d-h,m,n,o**) In contrast, other biomarker pairs, including CXCL10/CXCL9, PDGF-BB/EGF and CXCL9/ferritin exhibited correlations in both low-G and high-G (**Supplementary Figure 2b, c, l**). However, fibrinogen/ferritin and IL-6/Hb showed predominantly negative correlations in high-G (**Supplementary Figure 2i,p**). Therefore, these results indicated that a immunological markers are increased in patients with irCRS, they correlate with the severity of the clinical grading, and are positively or negatively correlated with conventional biological markers.

### Variations in immune cell subsets are associated with clinical severity of irCRS

Due to the limited sample size, we pooled mass cytometry data of patients into low-G (G1, G2) and high-G (G3, G4) for analysis. As we observed multiple differences in several immune subsets, we will only describe the most profound variations common to both low and high G irCRS (**Supplementary Figure 3, p values in Supplementary File 1**). The analysis of T-cell dynamics reveals a significant increase in several activated memory T cell subsets of CD4+ and CD8+ T cells, such as central memory (CM), transitional memory (TM), and effector memory (EM). These subsets were characterized by HLA-DR+ and CD38+ expression (**Supplementary Figure 3a-3b**). Additionally, we observed an increased frequency of CD38+ monocytes in classical (CL), intermediate (ITM), and non-classical memory (NCL), and a decrease in CD141+ DC, conventional mDC, and pDC (**Supplementary Figure 3c and 3e**). Mature neutrophils decreased while immature neutrophils and all CD38+ and CD62L+ neutrophil subsets increased (**Supplementary Figure 3f**). Other subsets showed no significant changes (**Supplementary Figure 3d-3g**). The only major difference between high and low-G was the decreased frequency of switched memory (SM) IgA2+ B cells and CXCR3+ NCL monocytes in low-G (**Supplementary Figure 3**).

### Identification of clusters with different survival linked to distinct cytokine profiles by using classical biomarkers

It was then determined whether distinct biomarkers profiles defined different survival groups based on their biomarkers profile. We thus performed K-means unsupervised clustering and several biomarkers were considered, including those used in the HScore, alone (**Fig. 2c**), combined with CRP (**Fig. 2d, Supplementary Table 4**) or creatinine (**Fig. 2e**) or both (**Fig. 2f, Supplementary Table 4**). To determine the optimal number of clusters, K-means clustering was evaluated for 1 to 4 clusters using the silhouette method. An example of the clustering performance on the PCA dimension-reduced plot for clusters C1 to C3 (**Fig. 3a**) and the corresponding silhouette curve (**Fig. 3b**) are shown. The HScore alone did not allowed identification of clusters associated with different survival from the onset of irCRS. However, the addition of biomarkers such as CRP and creatinine improved clustering performance. Using this approach, the most insightful results were obtained when multiple biological biomarkers were combined, including CRP, ferritin, creatinine, AST, ALT, total bilirubin, PAL, GGT, leukocytes, neutrophils, and Hb (**Fig. 3**). This approach led to the identification of three distinct patient clusters in a two-steps approach (**Fig. 3a-3b and Supplementary Table 5**). First, a cluster of three patients (Cluster 3) with significantly lower survival from irCRS onset and a combined cluster (1+2) were identified (**Fig. 3a, 3c**). On the remaining combined cluster (1+2), optimal clustering identified two distinct clusters, Cluster 1 and Cluster 2. Survival outcomes from the onset of irCRS differed significantly among the three clusters. Cluster 1 showed the most favorable prognosis, with a 100% 2-year survival rate, Cluster 2 presented an intermediate survival outcome, with 47% of patients surviving at 2 years and Cluster 3 had a 0% survival rate at 6 months (**Fig. 3c**). These survival trends were further supported by distinct profiles of overall survival (OS) and survival from the initiation of ICI treatment (**Fig. 3c**). Interestingly, the three clusters showed different patterns of clinical severity. Cluster 1 had a mixed composition of high-G cases, including both grade 3 (G3) and grade 4 (G4) irCRS cases whereas cluster 2 was predominantly low-G with 75% of cases. Notably, cluster 3 was characterized exclusively by the presence of three patients with irHLH thus confirming being the cluster associated with the most severe clinical presentation (**Fig. 3d**). Further delineation of clusters differences through biological profiling revealed that cluster 3 was also associated with the highest degree of biological perturbations and impact on multiple organs. Significant abnormalities were observed in markers of liver (AST, ALP) and kidney (creatinine) functions, of inflammatory markers (CRP, ferritin, lymphopenia), of coagulation (prothrombin ratio, activated partial thromboplastin time, d-dimer), and hematology (anemia) (**Fig. 3e-3h**).

**Figure 2.**
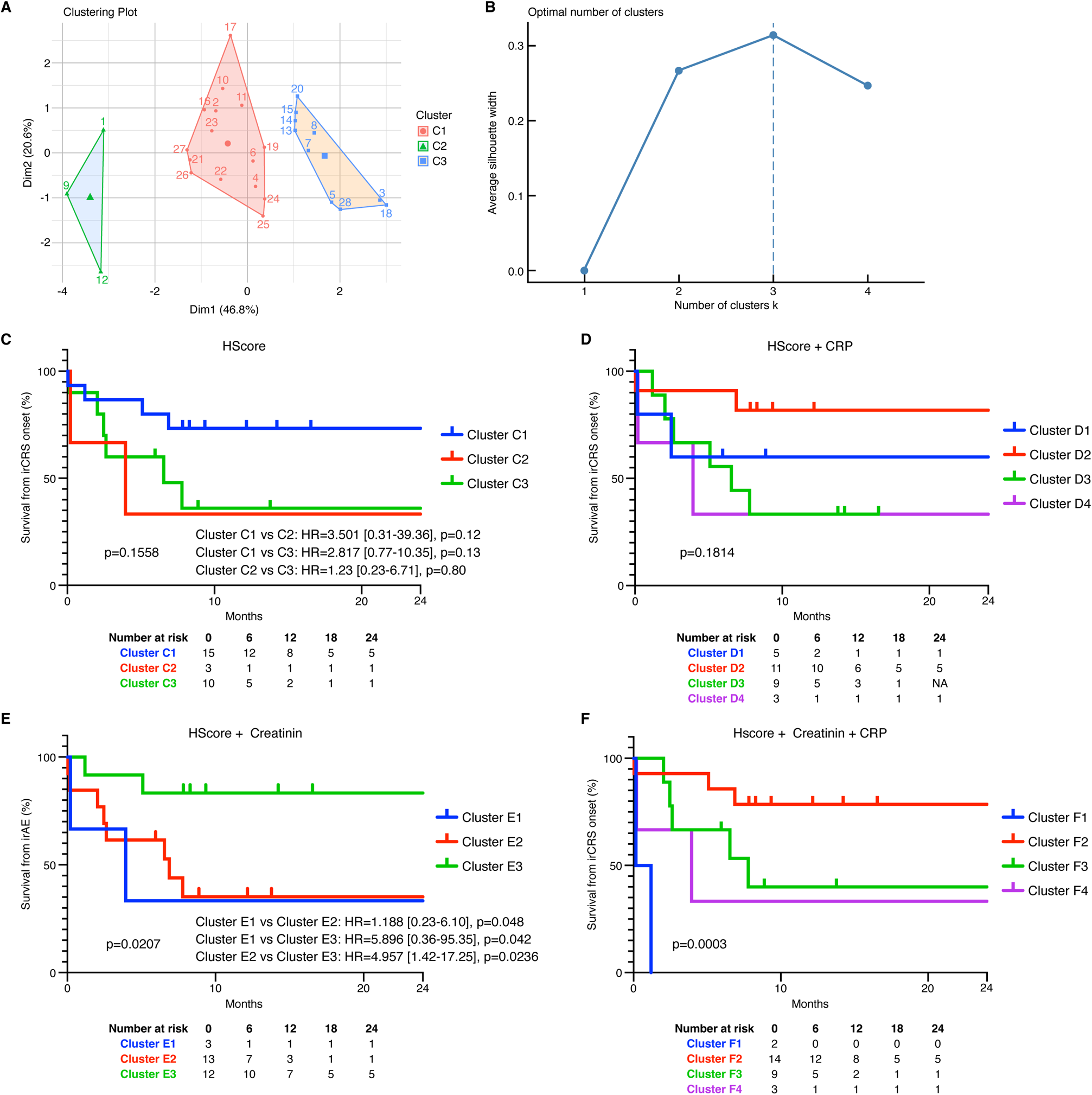
Exploration of 4 distinct clusterings across HScore and other biological parameters reveals variable strengths regarding survival from irAE. Biological parameters were collected from irCRS patients at the time of diagnosis (n=28). (**a**) clustering plot for the three identified clusters based on HScore parameters. (**b**) silhouette plot showing the optimal number of clusters based on HScore parameters. (**c**) Kaplan-Meier (KM) plot showing the survival rate from irCRS onset for the three Clusters (C1, C2, C3) obtained using HScore parameters. (**d**) KM plot showing the survival rate from irCRS onset for the four Clusters (D1, D2, D3, D4) obtained using a combination of HScore parameters and CRP. (**e**) KM plot showing the survival rate from irCRS onset for the three Clusters (E1, E2, E3) obtained for the four Cluster (F1, F2, F3, F4) using a combination of HScore parameters and creatinine. (**f**) KM plot showing the survival rate from irCRS onset using a combination of HScore parameters, creatinine, and CRP. Log-rank tests were used to analyze the data for statistical significance between groups and to calculate confidence intervals (CI) and hazard ratios (HR).

**Figure 3.**
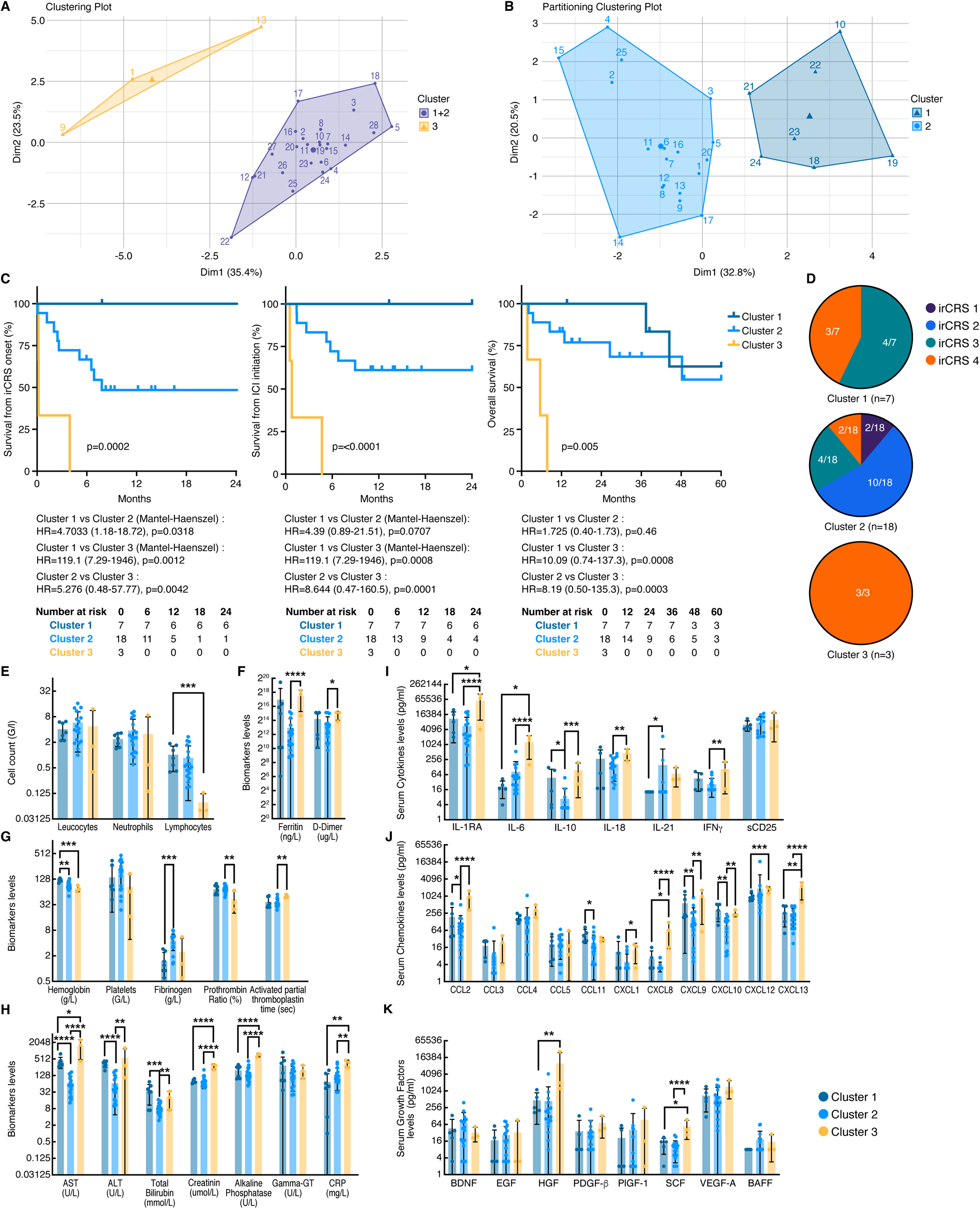
Integration of additional circulating biomarkers enhances clustering linked to distinct overall survival and cytokine profiles. The biomarkers used for this other clustering are CRP, ferritin, creatinine, AST, ALT, total bilirubin, ALP, GGT, leukocytes, PNN and Hb collected from irCRS patients at the time of diagnosis (n=28). (**a**) Clustering plot initially identified two distinct clusters: combined Cluster 1+2, and Cluster 3. (**b**) After isolating the three patients belonging to cluster 3, K means clustering was applied to the remaining cohort, resulting in the separation of the combined Cluster (1+2) into two new distinct clusters, labeled as Cluster 1 and Cluster 2 (**c**) Kaplan-Meier plot showing the survival rate for the three identified clusters, Cluster 1 (n=7), Cluster 2 (n=18) and Cluster 3 (n=3), the survival from irCRS onset, the overall survival (OS) or the survival from ICI initiation. (**d**) distribution of the irCRS according to the clinical grade in each cluster. (**e-h**) comparison of levels of circulating blood cells and serum biomarkers, including liver and kidney functions, coagulation and inflammation parameters in each cluster. (**i-k**) comparison of serum cytokine, chemokine, and growth factor levels among the three clusters. Log-rank or Mantel-Haenszel (if a cluster had 0 deaths during the interval) was used to calculate CI, HR, and p values for the KM curves. Student T tests were used to analyze the data for statistical significance of biomarkers levels between groups. Panels E to K plots represent values with individual data points, bar represent the mean and error bars represent standard deviation. CI, Confidence Interval; HR, Hazard Ratio, CRP: C-reactive protein; Hb: hemoglobin; PNN: polynuclear neutrophils; AST: aspartate aminotransferase; ALT: alanine transaminase; ALP: alkaline phosphatase; GGT: gamma glutamyl transferase.

Additional characterization through immune profiling revealed that Cluster 3 was characterized by a highly inflammatory profile with elevated levels of various cytokines (IL1-RA, IL-6, IL-10, IL-18, IFN-γ), chemokines (CCL2, CXCL8, CXCL9, CXCL10, CXCL12, CXCL13), and growth factors (HGF, SCF) (**Fig 3i-3k**). This inflammatory profile indicates a robust reactive inflammatory response, likely contributing to the clinical severity observed in Cluster 3. To assess the individual weight of different biomarkers on survival after the onset of irCRS, a 1D K-means clustering technique was applied to all measured cytokines and biological biomarkers, followed by univariate Cox proportional hazards modeling. As shown in **Supplementary Figure 4a-4b**, this methodological approach facilitated the identification of high and low clusters for each biomarker, including creatinine, hemoglobin, and prothrombin ratio. Analysis revealed that clusters characterized by high creatinine, low hemoglobin, or low prothrombin ratio were significantly correlated with reduced survival after irCRS onset. The results showed a hazard ratio (HR) of 5.605 (p=0.0068) for the high creatinine cluster, an HR of 6.643 (p=0.015) for the low hemoglobin cluster, and an HR of 5.811 (p=0.0324) for the low prothrombin ratio cluster (**Supplementary Figure 4a**). These findings were further supported by distinct Kaplan-Meier survival curves, showing the negative impact of these biomarker levels on patient outcomes post-irCRS onset (**Supplementary Figure 4c-4e**). Interestingly, when dual biomarkers such as hemoglobin and creatinine were combined for survival prediction, the accuracy was significantly improved. However, the predictive value reached its peak-100% accuracy-when hemoglobin, creatinine and prothrombin ratio were integrated into a unified clustering model (**Supplementary Figure 4f-4g**). This improvement in prognostic prediction by a multi-biomarker approach highlights the substantial benefits of including a combination of biological markers to assess survival outcomes in irCRS patients.

**Figure 4.**
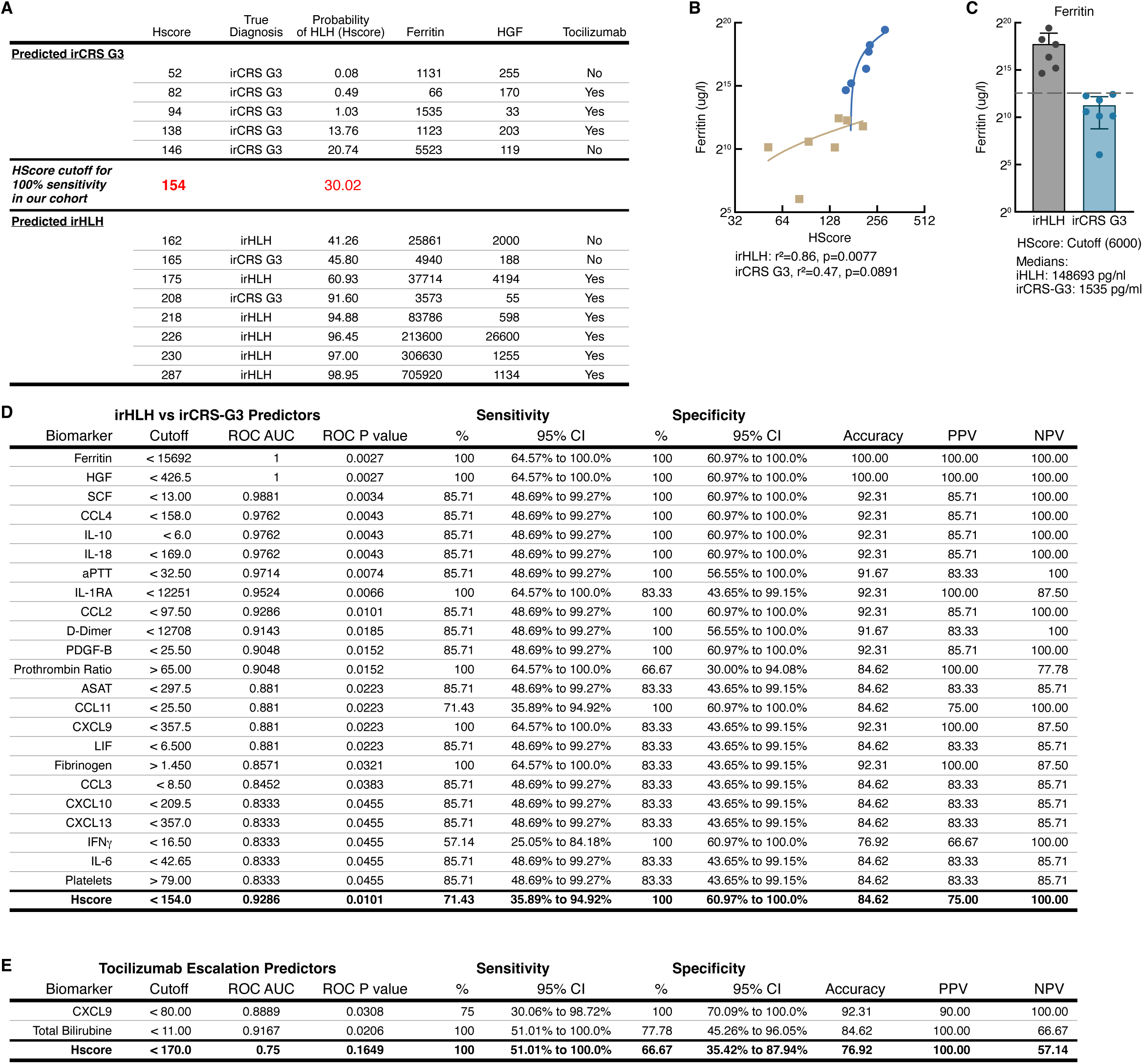
Superior performance of HGF and ferritin in discriminating between irCRS G3 and irHLH, with CXCL9 and total bilirubin predicting tocilizumab therapy escalation. (**a**) Comparison between the real diagnosis of high-G irCRS and estimation of diagnosis based to the conventional HScore. (**b**) Linear regression and correlation coefficient between ferritin and HScore in patients with irCRS G3 (n=7) or irHLH (n=6). (**c**) Median ferritin (pg/mL) in patients with irCRS G3 (n = 7) or irHLH (n = 6). (**d**) performance (area under the receiver operating curve (AUC), cut-off value, sensitivity, specificity, positive and negative predictive values) of each marker to discriminate irHLH (n=6) from irCRS G3 (n=7). (**e**) Performance (area under the receiver operating curve (AUC), cut-off value, sensitivity, specificity, positive and negative predictive values) of each marker in the prediction of treatment intensification with TCZ in patients with high-G irCRS (n=13). Confidence intervals for specificity and sensitivity have been calculated using Wilson-Brown method. Panel c plot represent values with individual data points, bar represent the mean and error bars represent standard deviation.

### Biomarker performance in differentiating irCRS Grade 3 from irHLH and predicting clinical severity and tocilizumab therapy escalation

As mentioned above, the primary objective was to identify biomarkers to improve diagnostic accuracy, the evaluation of clinical severity and to predict treatment intensification in patients with irCRS and irHLH. 45 serum biomarkers were evaluated including cytokines and biological markers in addition to those present in the traditional HScore(*31*). Our results confirmed the significant limitations of the HScore in the assessment of severe irCRS in patients with high-G irCRS. In fact, certain patients, despite low HScore, required escalating treatment with tocilizumab (TCZ), as observed in patients 2, 7, 9, and 11 (**Supplementary Figure 5a**). In irHLH patients, an HScore above 154 achieved 100% diagnostic sensitivity, identifying 2 patients with low HScore HLH probability (41% and 61%) (*31*) (**Fig. 4a**). In addition, a strong correlation was found between higher ferritin levels and increased clinical severity in patients with irHLH (**Fig. 4b**), with median ferritin levels > 148000 in the irHLH group well above the established highest HScore cutoff of 6000 (**Fig. 4c**). These results highlight the limitations of the traditional ferritin threshold of 6000 of the HScore (*31*) in this context, which adds only 50 points and thus fails to capture the full extent of the inflammatory state associated with irHLH, where ferritin could rise to much higher levels.

Based on these limitations, we attempted to identify the most reliable predictive biomarkers differentiating between irHLH and irCRS G3. As detailed in the demographics, all the irCRS G4 were irHLH. Of the irCRS G3, one patient was irHLH but was excluded from the biomarker analysis because the biomarker panel was performed after initiation of immunosuppressive treatment. We analyzed 45 biomarkers including, 27 cytokines, 18 biological markers including those of the HScore. We determined the cutoff values, area under the receiver operating characteristic curve (AUC), sensitivity, specificity, positive and negative predictive values, and accuracy. This analysis identified a panel of 24 biomarkers with P-values below 0.05, ranging from <0.0455 to <0.0027 (HGF, ferritin, SCF, IL-10, IL-18, CCL4, aPTT, IL-1RA, CCL2, D-dimer, PDGF-BB, CXCL8, prothrombin time ratio, CXCL9, CCL11, LIF, ASAT, CXCL10, fibrinogen, CCL3, IL-6, CXCL13, IFN-γ and platelets) that significantly discriminated between irHLH and irCRS G3 **(Fig 4d).** Among them, HGF, ferritin, IL-1RA, prothrombin time ratio, CXCL9 and fibrinogen showed the highest sensitivity (100%) with specificity ranging from 66% to 100%. Remarkably, HGF and ferritin showed excellent 100% accuracy whereas the sensitivitythat of the HScore was only 71.43% **(Fig 4d)**. Importantly, only CXCL9 and total bilirubin were significantly predictive of treatment intensification with TCZ, with sensitivities of 75 (CXCL9) to 100% (bilirubin) and specificities of 77.78% (bilirubin) to 100% (CXCL9) whereas the specificity of HScore was 66.67% **(Fig 4e)**. Regarding hemodynamic instability requiring fluid resuscitation, HGF, SCF and aPTT were the best predictors with 100% accuracy, whereas the accuracy of the HScore was only 76.92% (**Supplementary Figure 5b**). For respiratory distress, CCL2, ferritin, D-dimer and aPTT were the only markers with 100% positive predictive value and sensitivity, with specificity ranging between 71.43-85.71%, wheras the HScore had 83.3% accuracy and 85.71% positive predictive value (**Supplementary Figure 5c**).

### High-grade irCRS and sepsis patients exhibit distinct biomarker profiles

Although patients with high-G irCRS and sepsis may present with similar clinical manifestations, it is crucial to differentiate between these two conditions. For these purposes, we compared the biomarker levels of patients with irCRS G3 or irHLH to those of patients with sepsis and identified a cytokine and immune cell signature that distinguished them. The patients with irHLH had significantly higher levels of IL-10 and IL-1RA (**Supplementary Figure 6a and 6d**). sCD25 was not significantly different between groups (p=0.08), but IL-6 was highest in sepsis compared to irCRS G3 (**Supplementary Figure 6a, 6d**). IFN-γ pathway was observed in irHLH, as indicated by significant increases in IFN-γ, CXCL-9, CXCL-10 and IL-18, as well as CCL3 and CCL4, compared to sepsis and irCRS G3 (**Supplementary Figure 6a and 6b**). IL-7 was significantly increased in sepsis (**Supplementary Figure 6a**). HGF was higher in irHLH compared to sepsis and irCRS G3, whereas EGF and GM-CSF were higher in sepsis compared to irCRS G3 (**Supplementary Figure 6c**). SCF and LIF were higher in irHLH compared to irCRS G3 (**Supplementary Figure 6d**). Leukocytes and neutrophils were higher in sepsis compared to irHLH and irCRS G3 (**Supplementary Figure 6e**). Ferritin, D-dimer, and transaminases were more elevated in irHLH than in sepsis and irCRS G3 (**Supplementary Figure 6f**). Furthermore, irHLH showed more severe coagulopathies, including prothrombin ratio and fibrinogen levels (**Supplementary Figure 6g**). Mass cytometry analysis between high-grade irCRS (irHLH and G3) and sepsis (**Supplementary Figure 7, p values in Supplementary File 2**), identified CD38+ HLA-DR+ double-positive T cells as increased in the CM, EM, and TM subsets. Compared to sepsis, there was a significant increasing trend of CD4 memory CD38+, CD4 TM HLA-DR+/CD38+, CD8 memory CCR4+, and a decrease in CD8 memory CXCR5+ in high-grade irCRS. (**Supplementary Figure 7a and 7b**). Compared to sepsis, there was a significant increase in the frequency of CD38+ monocytes in the ITM subset. Additionally, there was a significant increasing trend in CL monocytes CD11c+ and CL monocytes CD38+ and in the frequency of CD16 NK cell (**Supplementary Figure 7d-7e**). Compared to sepsis, high-G irCRS exhibited decreased total neutrophils but increased CD62L neutrophils in both immature and mature subsets (**Supplementary Figure 7f**). No significant differences were observed in the other subsets (**Supplementary Figure 7d, 7e and 7g**).

### Discriminating irCRS Grade 3, irHLH, and sepsis through circulating biomarkers

The potential of all biomarkers to discriminate between irCRS grade 3, irHLH and sepsis at the time of diagnosis was evaluated. Multiple supervised approaches were used to evaluate this In the first approach, we analyzed 46 biomarkers including 27 cytokines, and 18 biological markers. We determined cutoff values, area under the receiver operating characteristic curve (AUC), sensitivity, specificity, positive and negative predictive values, and Youden’s J index for each biomarker. This analysis identified a panel of 11 biomarkers (leucocyte count, IL-7, fibrinogen, EGF, CXCL10, neutrophils, GM-CSF, ALAT, ALP, GGT and IL-6) that significantly discriminated between sepsis and irCRS high-G with P-values ranging from <0.0046 to <0.0415 **(**Fig. 5a**).** Among the biomarkers tested, leukocyte count, IL-7, fibrinogen, EGF, CXCL10, and neutrophils demonstrated the highest sensitivity (100%) with an excellent specificity of 92.31% (**Fig. 5a**). In addition, a panel of 10 biomarkers (leucocyte count, neutrophils, IL-6, EGF, ALP, IL-7, GM-CSF, fibrinogen, IL-15 and CXCL10) was identified and significantly discriminated between sepsis and irCRS G3 with P-values ranging from <0.0082 to <0.0472 (**Fig. 5b**). Among the measured parameters, leukocyte count, neutrophils, IL-6, and EGF exhibited an accuracy of 100% (**Fig. 5b**). In addition, the ferritin cut-off of 23221 ng/ml was discriminating between irHLH and the rest of patients (sepsis and irCRS G3) with sensitivity and specificity of 100 % (ROC p value of 0.0009) (**Fig. 5c and Supplementary Table 6)**. Two additional supervised approaches were used. First using sPLS-DA (Sparse Partial Least Squares Discriminant Analysis), we found that the biomarker profiles could effectively separate the three conditions, with only one irCRS case falling between the irCRS and Sepsis clusters (**Supplementary Figure 8a**). sPLS-DA first component identified 5 biomarkers with absolute importance higher than 0.3: IL-10, HGF, IL-18, IL-1RA, and SCF (**Supplementary Figure 8b**). Component 2 identified three variables exceeding 0.3: leukocytes count, IL-7, and EGF (**Supplementary Figure 8c**). Component 1 explained most of the X axis variation notably the separation of irHLH patients identifying biomarkers previously found using the ROC curve approach (*i.e* IL-10, HGF) and component 2 the Y axis separating the sepsis from the irHLH identifying also biomarkers identified in Figure 5 (*i.e* leucocytes count, IL-7, EGF). The potential for developing a decision tree construction was explored using the random forest algorithm to identify the most predictive biomarkers for classification. Ferritin emerged as the most abundant initial node in the decision trees, with ferritin, HGF, and leukocyte count ranking as the top three biomarkers based on their average Gini coefficient across 100 trees (**Supplementary Figure 8d and 8e**).

**Figure 5.**
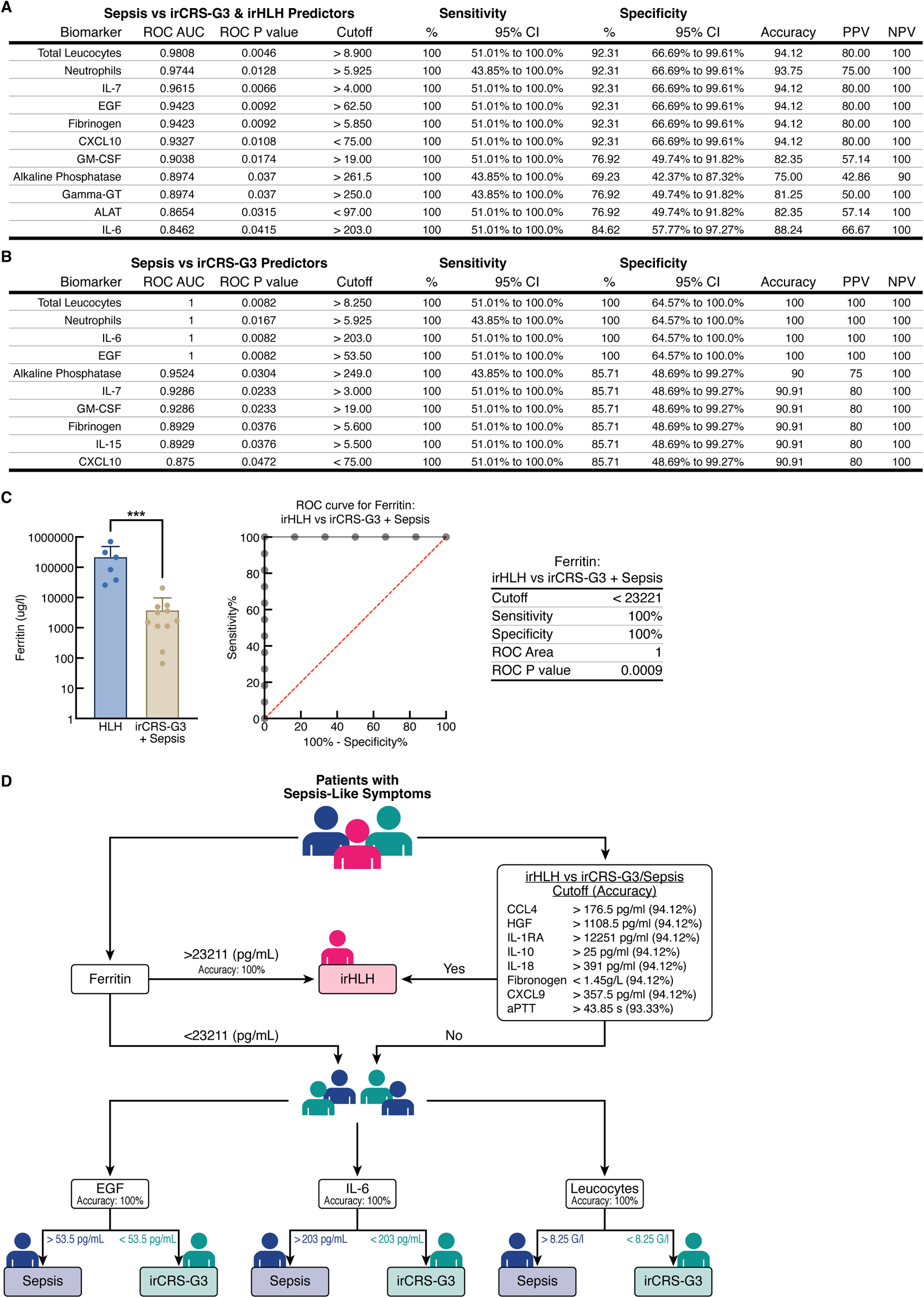
Distinct biomarker profiles in irCRS G3, irHLH and sepsis allow differential diagnosis. (**a**) Performance (area under the receiver operating curve (AUC), cut-off value, sensitivity, specificity, positive and negative predictive values) of each marker to discriminate sepsis (n=4) from high-G irCRS (n=13). (**b**) Performance of each marker to discriminate sepsis (n=4) from irCRS G3 (n=7). (**c**) ferritin levels in patients with irHLH (n=6) vs irCRS G3 and sepsis (n=4) with area under the receiver operating curve (AUC), cut-off value, sensitivity, specificity. (**d**) Decision tree analysis showing the performance of each biomarker in discriminating between irCRS G3, irHLH and sepsis. Confidence intervals for specificity and sensitivity have been calculated using Wilson-Brown method. Of note, neutrophil count, alkaline phosphatase, and gamma-GT were only performed in 3 sepsis patients because one patient had missing measurements at the time of inclusion. Panel c plot represent values with individual data points, bar represent the mean and error bars represent standard deviation.

Our aim was to develop a decision tree capable of distinguishing the three CRS-like presentations within our cohort. Using ferritin with a threshold of >23111 ug/L, 100% accuracy was achieved in discriminating irHLH patients from those with irCRS G3 or sepsis. Eight additional markers (CCL4, HGF, IL-1RA, IL-10, IL-18, fibrinogen, CXCL9 and aPTT) have higher than 90% accuracy to identify irHLH patients from irCRS-G or sepsis (**Supplementary Table 6**). The differentiation between irCRS G3 and sepsis was further refined using three biomarkers-EGF, IL-6, and leukocytes-with specific thresholds, EGF (>51.3 pg/mL), IL-6 (>129.6 pg/mL), and leukocytes (>8.5 G/L), that allowed perfect separation between these conditions. The integration of these biomarkers facilitated the prediction of irHLH, irCRS G3, and sepsis with 100% accuracy within our cohort (**Figure 5d**). Neutrophils count also have a 100% accuracy **(Figure 5b)** but was not included in the tree due to the lack of measurement in 1 sepsis patients. This multifaceted biomarker analysis underscores the effectiveness of integrating multiple immune profiling techniques to precisely distinguish between irCRS, irHLH and sepsis, which is critical for guiding appropriate therapeutic interventions in these clinically challenging scenarios.

### Tocilizumab is effective treatment of corticosteroid refractory high-grade irCRS

Twelve patients with severe high-G irCRS received adjunctive therapy with TCZ, an anti-IL-6R antagonist, due to a suboptimal initial response to corticosteroids. TCZ use in irCRS/irHLH patients adheres to our standard care in combination where it is employed as a second-line escalation therapy in corticosteroid-sparing strategies, or as an initial combination therapy with corticosteroids in severe CRS cases. This in accordance to our previous work (*15*) which contributed to our involvement in developing the irHLH ESMO guidelines(*1*). These patients’ data were retrospectively collected, and out of these 12 patients, 3 were not included in the rest of the manuscript as their cytokines were only collected after TCZ initiation. Encouragingly, a 100% response rate to treatment with TCZ was observed, resulting in rapid and substantial clinical improvements. Importantly, all cases of irHLH were resolved and no irCRS-related mortality was observed at either 7 or 30 day (**Supplementary Table 2**). Longitudinal cytokine follow-up data were available for only 6 out of 12 patients. In this subgroup, standard markers of inflammation such as cytolysis, ferritin levels, C-reactive protein (CRP) and leukopenia showed significant reductions and normalization (**Fig. 6a**). Similarly, TCZ treatment induced significant reductions in all immunological markers including IFN-γ, soluble CD25 (sCD25), IL-6, IL-18, CXCL9, CXCL10, CCL2, CCL4, CCL5, HGF, SCF, IL-10 and IL-1RA (**Fig. 6b)**. Importantly, TCZ therapy did not result in any significant adverse events.

**Figure 6.**
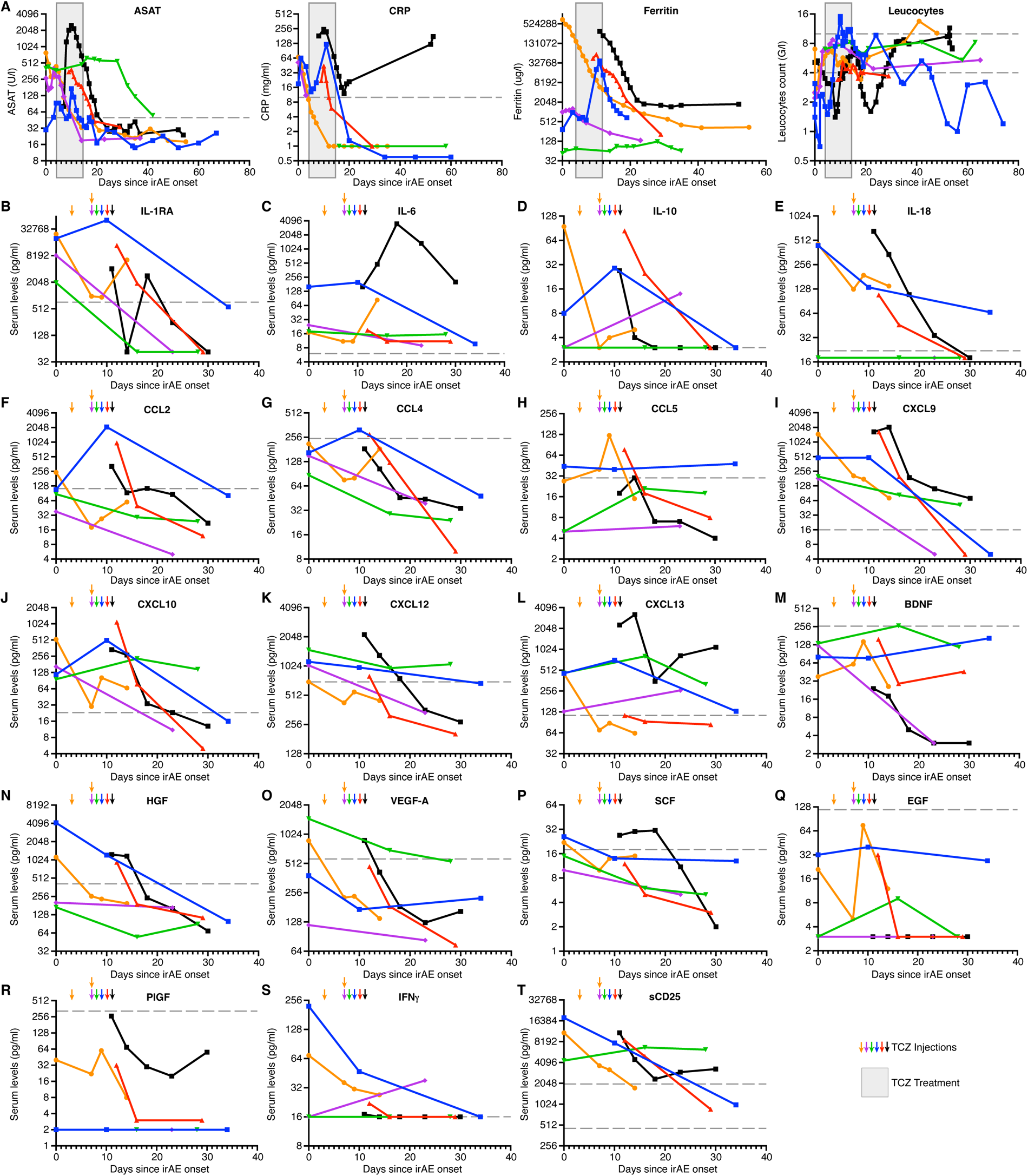
Longitudinal biomarker changes and biological resolution in CS-Refractory High-G irCRS patients treated with tocilizumab. (**a**) Evolution of serum biomarker levels including AST (UI/L), CRP (mg/L), ferritin (µg/L), leukocytes (G/L), and (**b**) serum cytokine, chemokine, and growth factor levels, in patients with repeated cytokines measurements receiving tocilizumab treatment for CS-refractory high-grade irCRS (n=6). The gray boxes in panel a indicate the global period of tocilizumab treatment, while the arrows in panel **b** indicate individual days of tocilizumab administration.

## Figures Legends

**Supplementary Figure 1:**
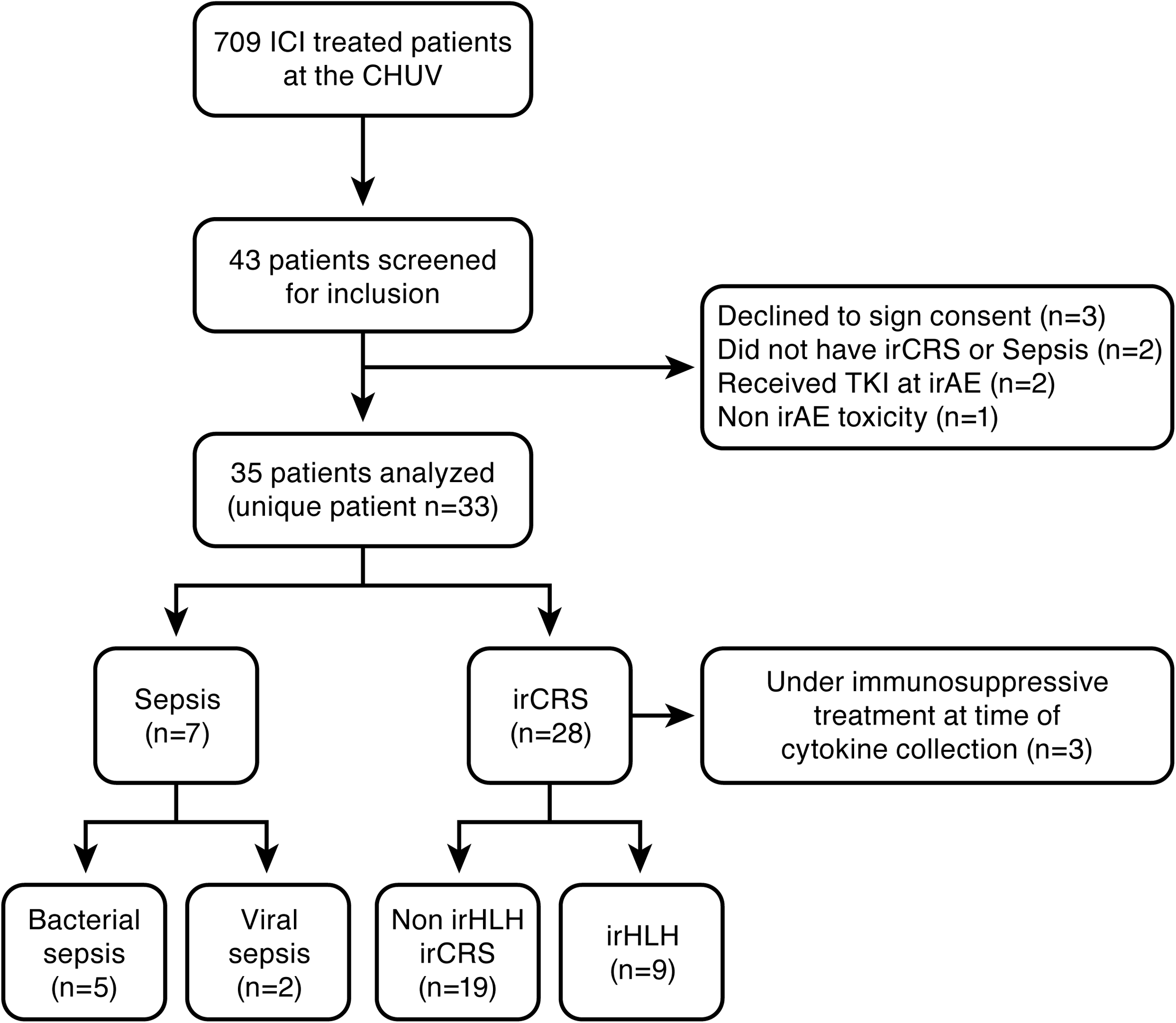
Study flowchart.

**Supplementary Figure 2:**
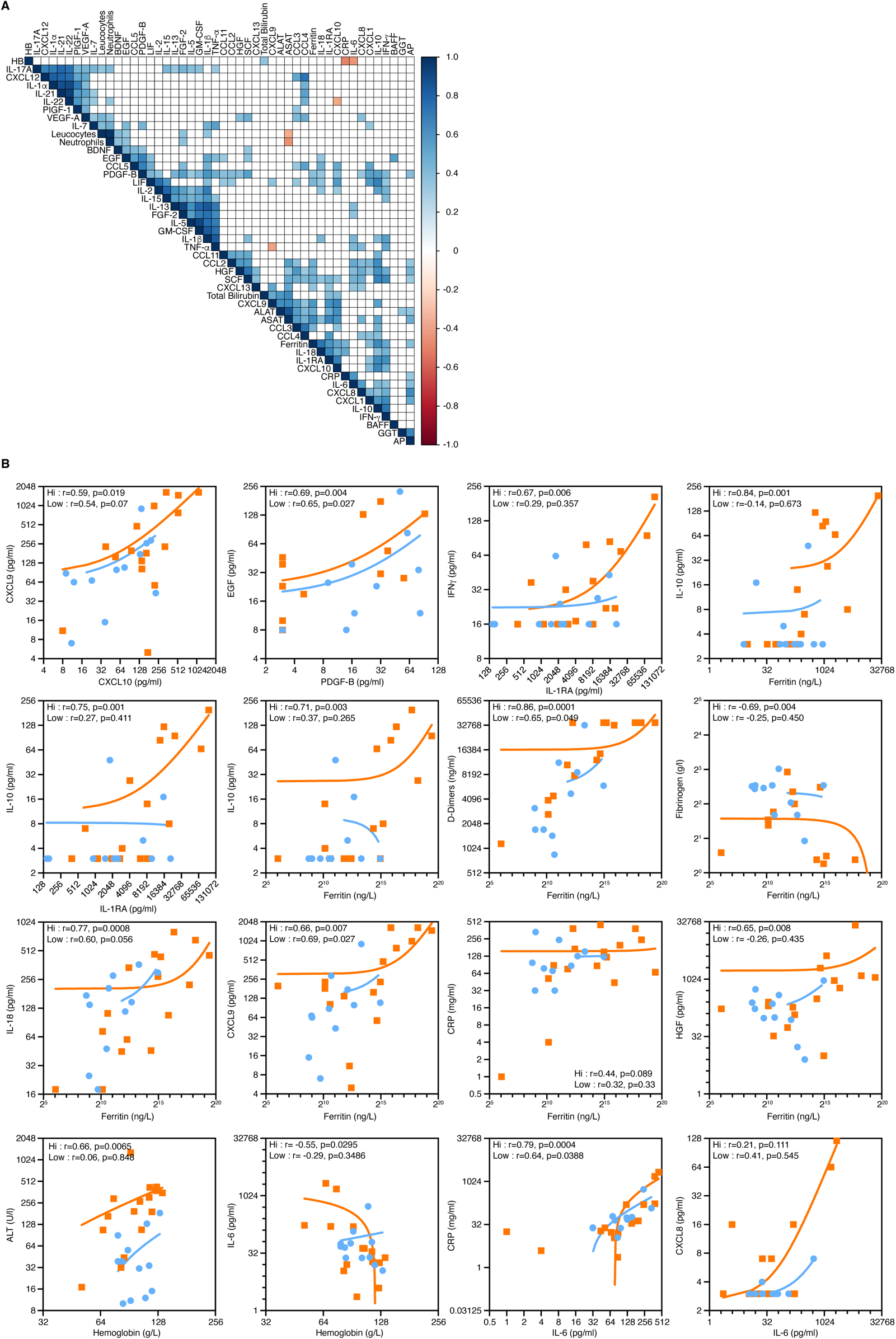
Correlation between cytokine and biological biomarkers by clinical grade of irCRS. **(a)** Spearman Correlation Matrix. Correlation map plotted using significance levels for the Spearman test performed with relevant serum biomarker data from all irCRS patients studied across all grades. Positive correlations are shown in shaded blue and negative correlations are shown in shaded red. Correlations with a p-value ≥ 0.05 are not considered significant and are left blank. Color intensities are proportional to the correlation coefficients. On the right side of the correlogram, the color legend shows the correlation coefficients and corresponding colors. (**b-q**) Correlation and linear regression between the level of various serum biomarkers including cytokines and biological parameters according to the clinical severity, low-grade (G1, G2) (n=12) in orange versus high-grade (G3, G4) (n=16) in blue. Each point represents a unique patient time point at irCRS diagnosis. Spearman’s rank correlation coefficient and significance are shown.

**Supplementary Figure 3:**
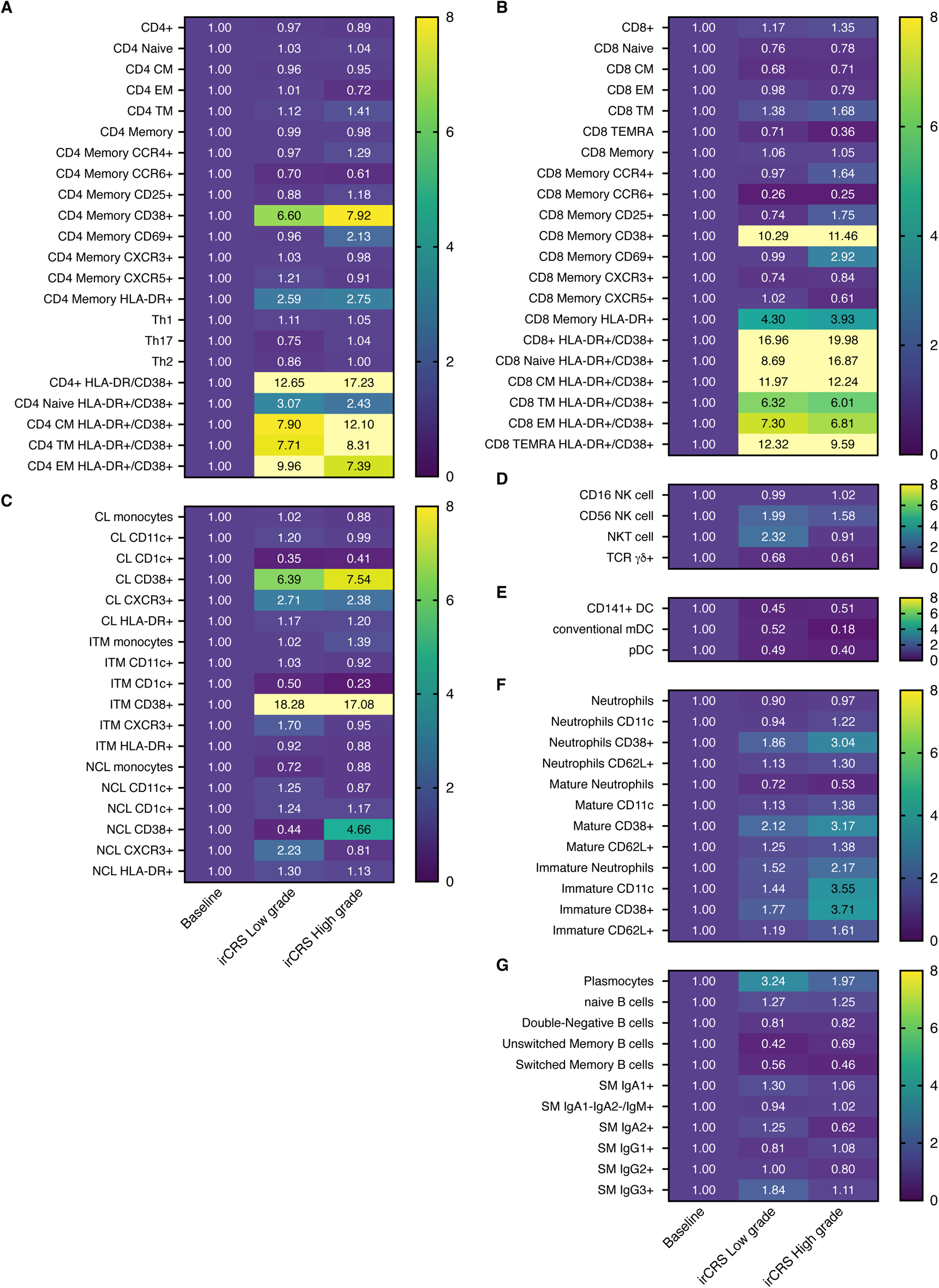
Comprehensive profiling of circulating immune cell phenotypes differentiates high from low irCRS grades. (**a-g**) Heatmap showing fold change relative to pre-ICI baseline of cancer patients (n=103) of the different immune cell subsets in patients at the time diagnosis of low-G irCRS (n=10) vs high-G irCRS (n=10). (**a**) CD4 T cell subsets, (**b**) CD8 T cell subsets, (**c-e**) monocyte subsets, (**f**) NK cells, NKT cells and TCRγδ T cells, (**g**) dendritic cells DCs, (**h**) neutrophils, (**i**) B cells. Values represent % of parent population as indicated in the gating strategy (Supplementary_OLD Figure 2). Statistical significance was analyzed using Student T test between groups. ***P<0.001, ****P<0. EM: effector memory, CM: central memory, DC: dendritic cells, TEMRA: terminally differentiated effector memory cells re-expressing CD45RA, classical monocytes CL, intermediate monocytes ITM, and non-classical monocytes NCL

**Supplementary Figure 4:**
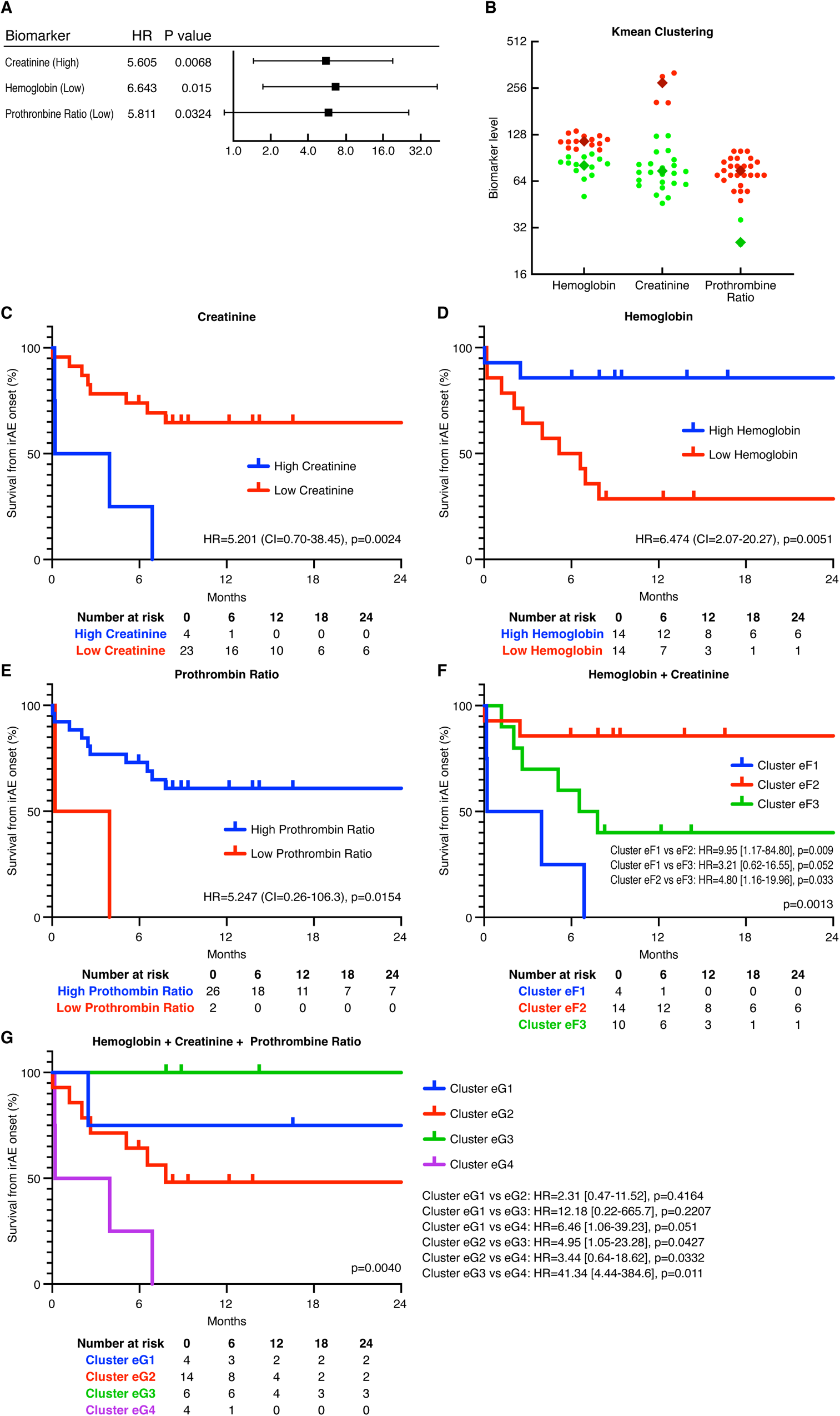
1d K-Means clustering of biological parameters reveals high and low clusters with differential impact on survival from irAEs. (**a**) Univariate Cox regression models for OS with the different high and low clusters identified by K-means clustering for various biomarkers (n=28). (**b**) Biomarkers levels in the different identified low and high clusters. (**c**) Kaplan-Meier curves illustrating the survival from irCRS onset in low (n=4) and high (n=23) creatinine clusters. (**d**) Survival in low (n=14) and high (n=14) hemoglobin clusters. (**e**) Survival in low (n=2) and high (n=26) prothrombin ratio clusters. (**f**) Kaplan-Meier plot showing the survival rate for the three Clusters, Cluster eF1 (n=4), Cluster eF2 (n=14) and Cluster eF3 (n=10), identified with K-means using hemoglobin (Hb) and creatinine, (**g**) Kaplan-Meier plot showing the survival rate for the four clusters, Cluster eG1 (n=4), Cluster eG2 (n=14), Cluster eG3 (n=6) and Cluster eG4 (n=4) identified with K-means using creatinine, Hb and Prothrombin ratio. Log-rank (Mantel-Cox) test was used to identify significant differences between groups in panels F and g, logrank for comparing two clusters in c to e.

**Supplementary Figure 5:**
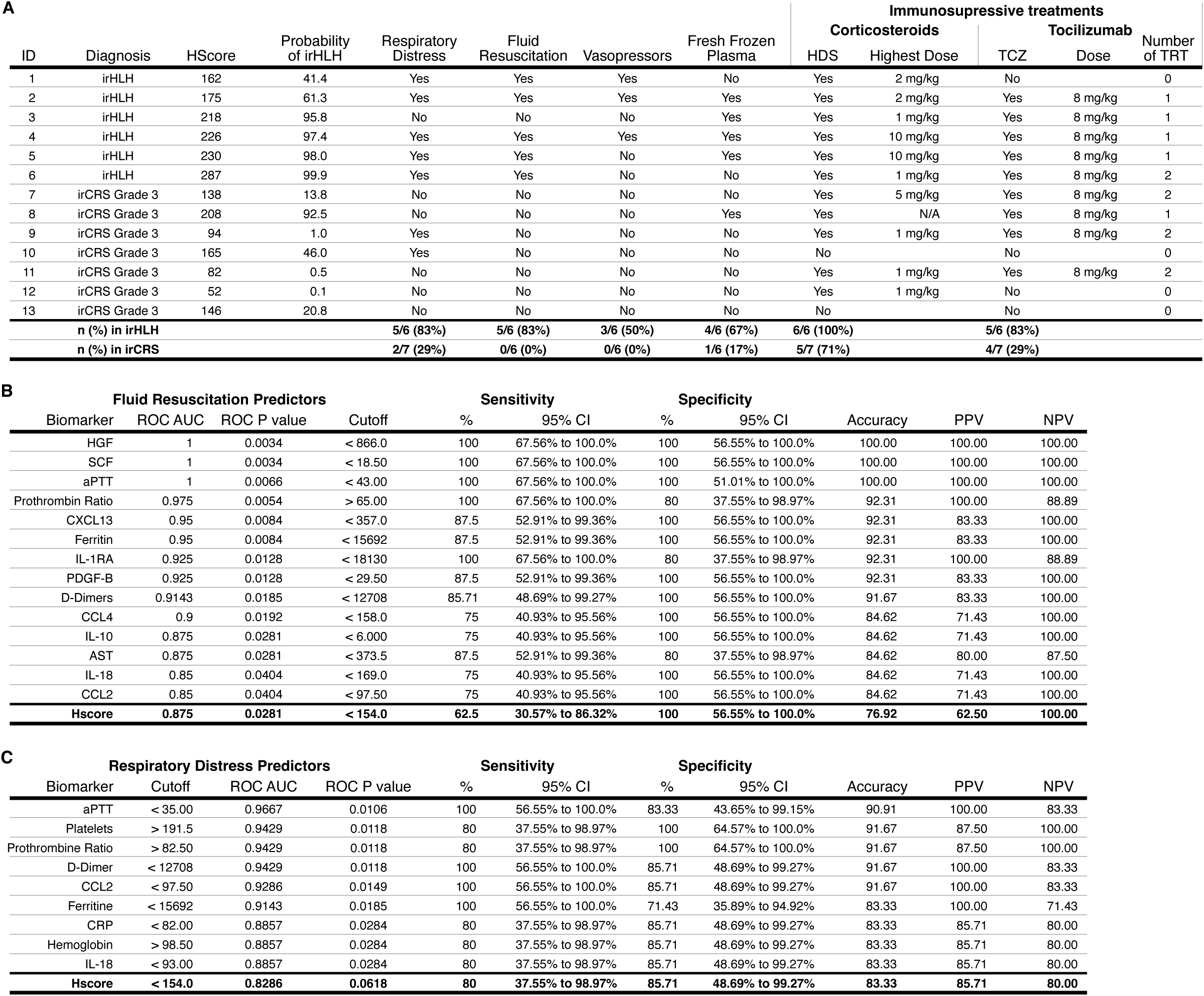
Biomarker efficacy in the prediction of hemodynamic instability and respiratory distress in patients with high-G irCRS. **(a)** clinical and treatment characteristics of all patients with high-G irCRS (n=13). (**b**) Performance (area under the receiver operating curve (AUC), cut-off value, sensitivity, specificity, positive and negative predictive values) of each marker in the prediction of hemodynamic instability requiring vasopressor in patients with high-G irCRS (n=7). (**c**) performance (area under the receiver operating curve (AUC), cut-off value, sensitivity, specificity, positive and negative predictive values) of each marker in the prediction of respiratory distress in patients with high-G irCRS (n=13). Confidence intervals for specificity and sensitivity have been calculated using Wilson-Brown method. TCZ: tocilizumab

**Supplementary Figure 6:**
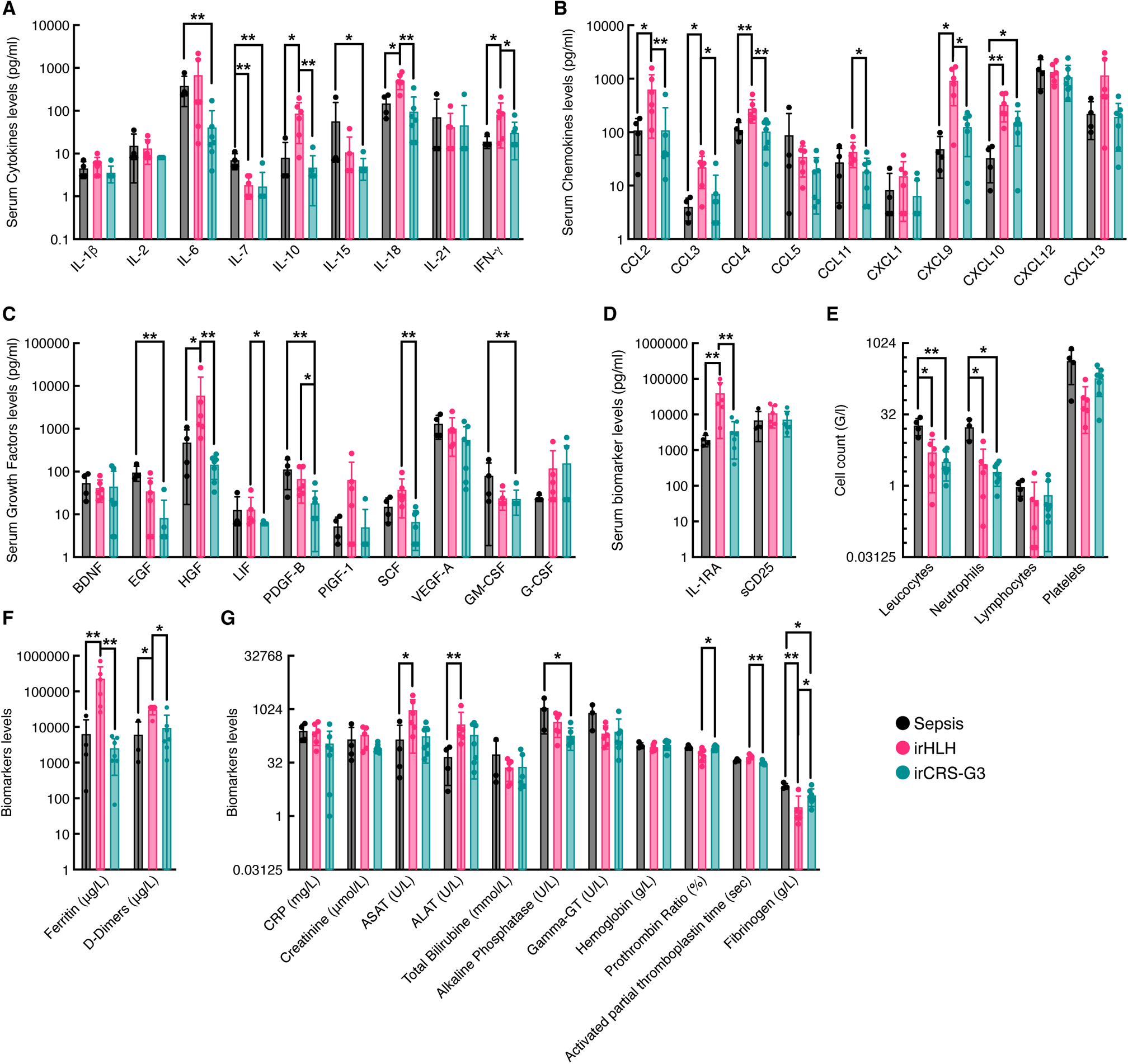
Cytokine and Biological profiles for irCRS G3, irHLH and Sepsis. (**a-d**) comparison of serum cytokine, chemokine, and growth factor levels. (**e-g)** circulating blood cells and serum biomarker levels (including liver and kidney functions, coagulation and inflammation parameters) in the three groups of patients: irCRS G3 (n=7), irHLH (n=6) and sepsis (n=4). Panels A to G plots represent values with individual data points, bar represent the mean and error bars represent standard deviation. Statistical significance was analyzed using Mann-Whitney test between groups. ***P<0.001, ****P<0.0001.

**Supplementary Figure 7:**
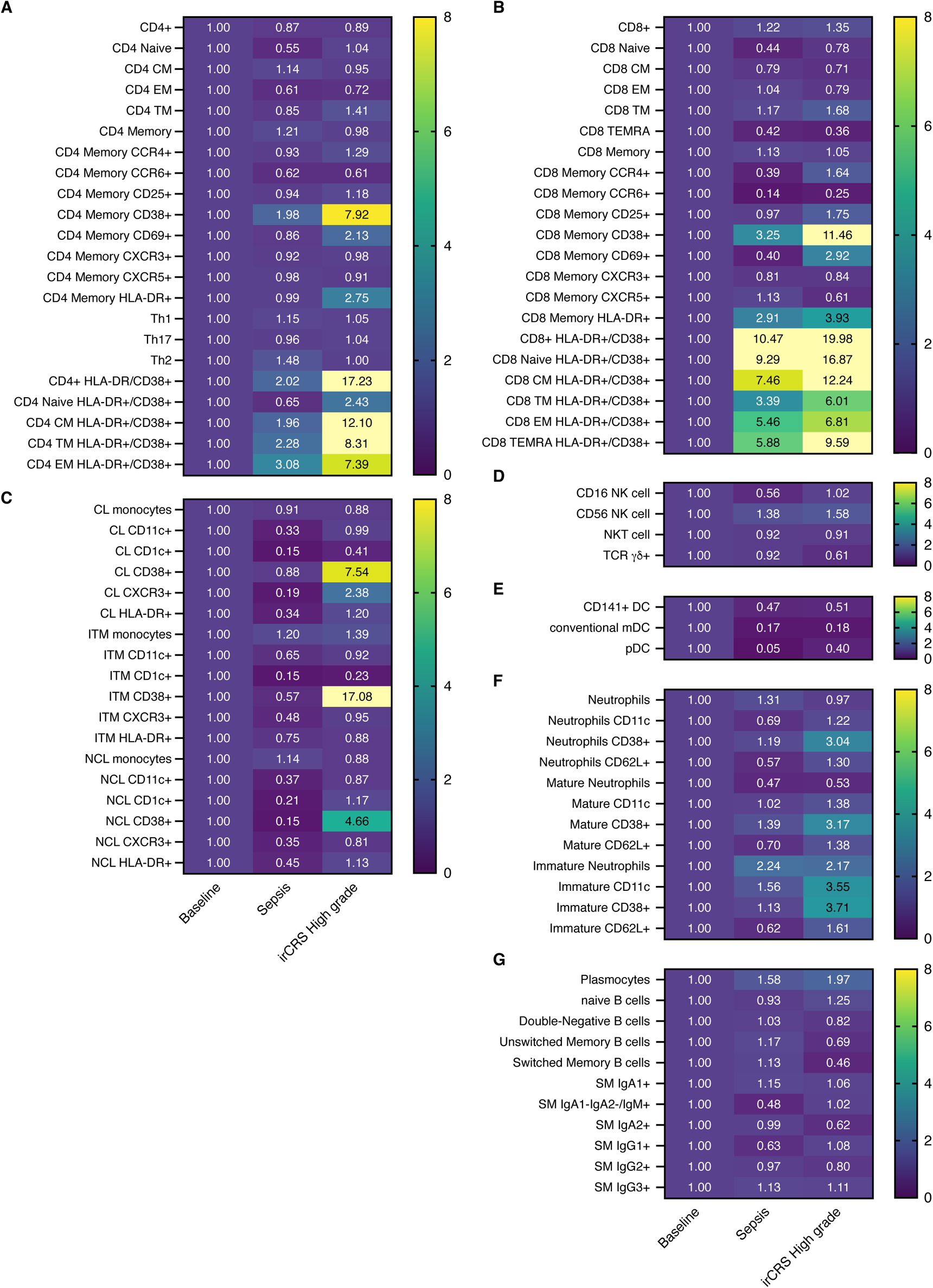
Comparative distribution of circulating immune cell populations in patients with irCRS G3, irHLH or sepsis. (**a-g**) Heatmap showing fold change relative to pre-ICI baseline of cancer patients (n=103) of the different immune cell subsets in patients with irCRS G3 (n=4), irHLH (n=6) or sepsis (n=5). (**a**) CD4 T cell subsets, (**b**) CD8 T cell subsets, (**c-e**) monocyte subsets, (**f**) NK cells, NKT cells and TCRγδ T cells, (**g**) dendritic cells DCs, (**h**) neutrophils, (**i**) B cells. Values represent % of parent population as indicated in the gating strategy (Supplementary_OLD Figure 2). Statistical significance was analyzed Student T tests between groups. ***P<0.001, ****P<0.0001. EM: effector memory, CM: central memory, DC: dendritic cells, TEMRA: terminally differentiated effector memory cells re-expressing CD45RA, classical monocytes CL, intermediate monocytes ITM, and non-classical monocytes NCL

**Supplementary Figure 8:**
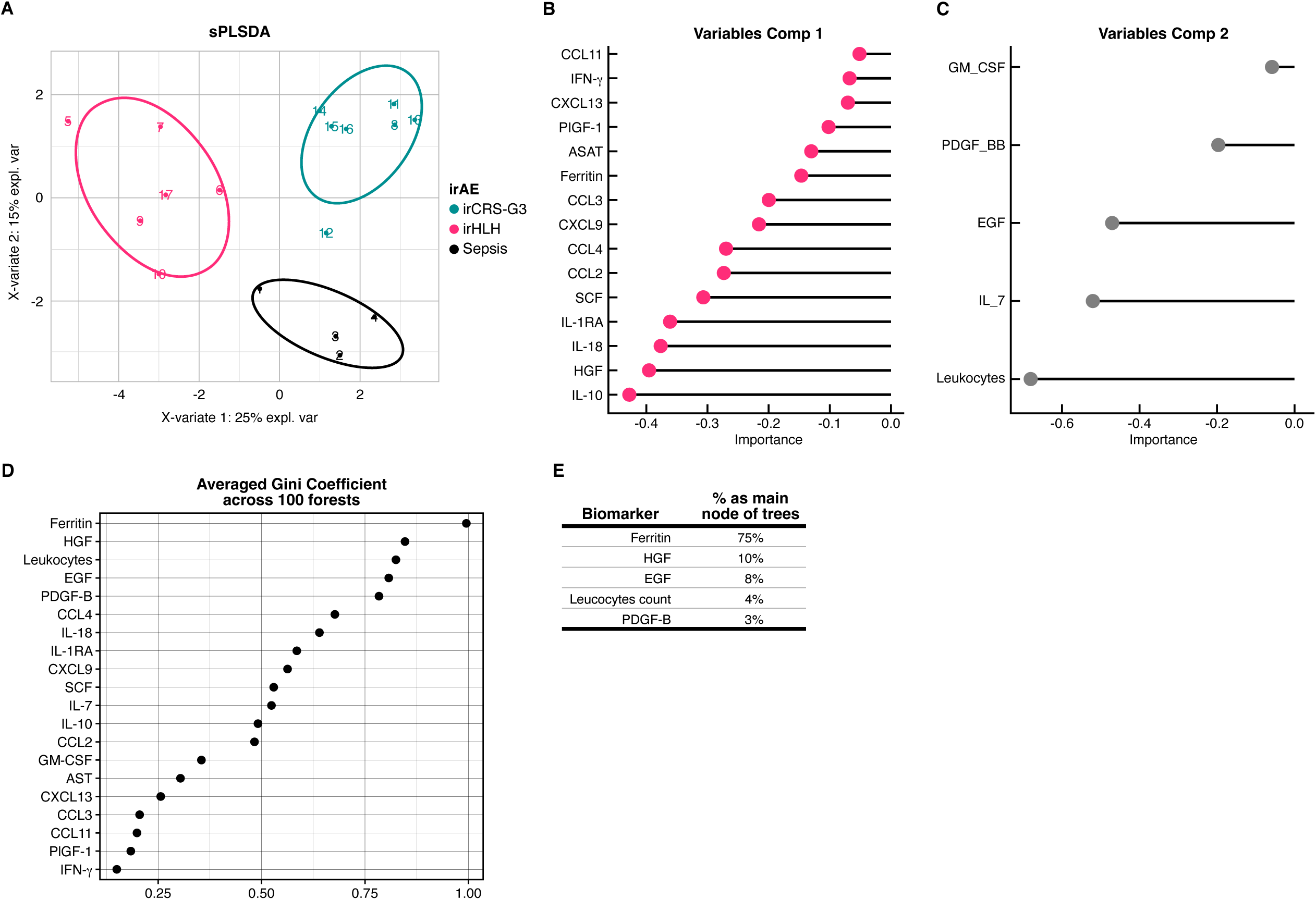
Clustering of circulating biomarkers provides distinction between irCRS, irHLH and sepsis. (**a**) Supervised learning sPLS-DA approach used to separate the three subgroups: irCRS G3, irHLH and sepsis. (**b**) The variables of component 1 allow for the separation of irHLH from the other 2 groups. (**c**) The variables of component 2 allow the separation of irCRS grade 3 from sepsis. (**d**) random forest averaged Gini coefficient across 100 forest for all biomarkers. (e) Percentage as the main node of trees for all biomarkers. sPLS-DA: sparse partial least squares discriminant analysis.

**Supplementary Figure 9:**
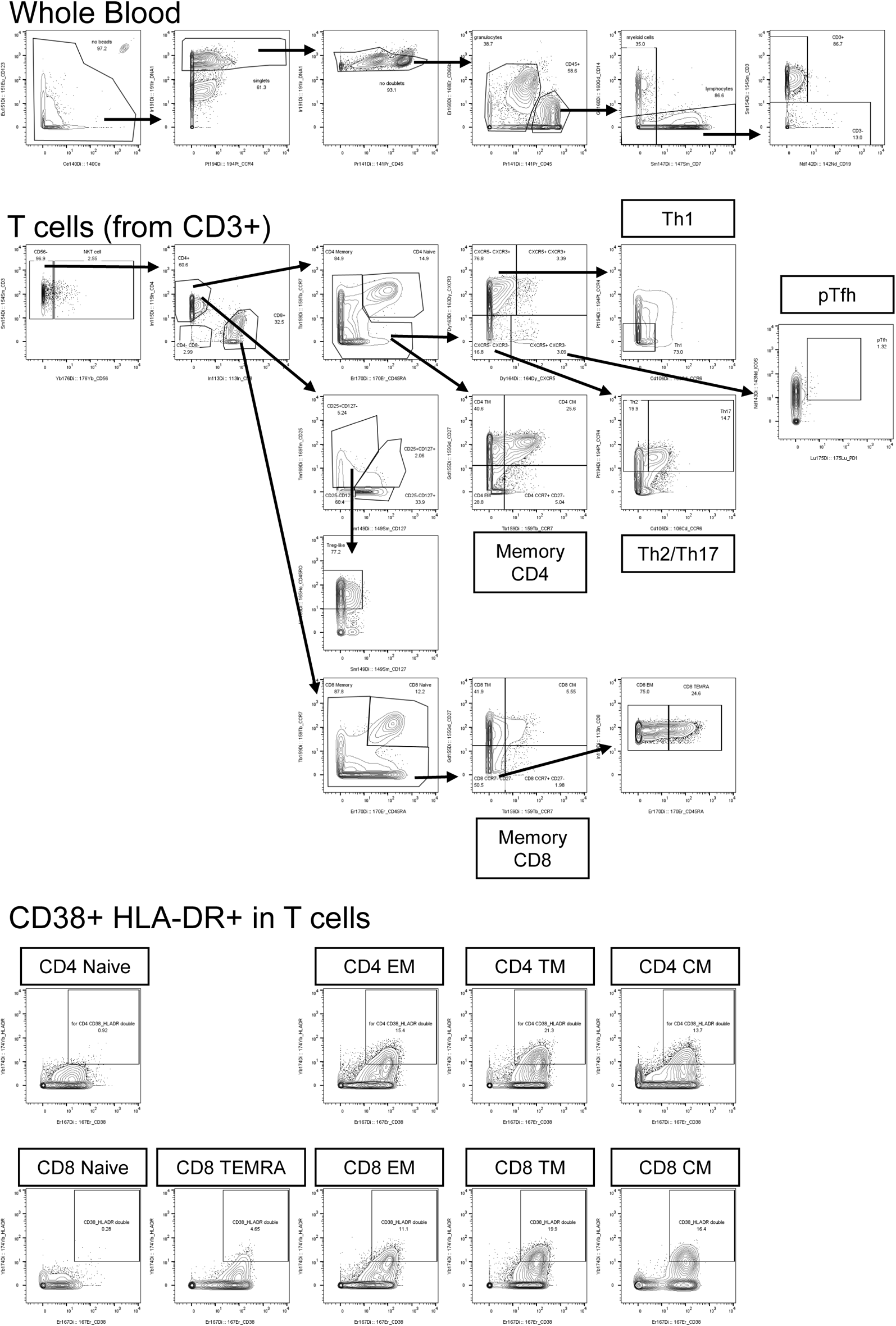
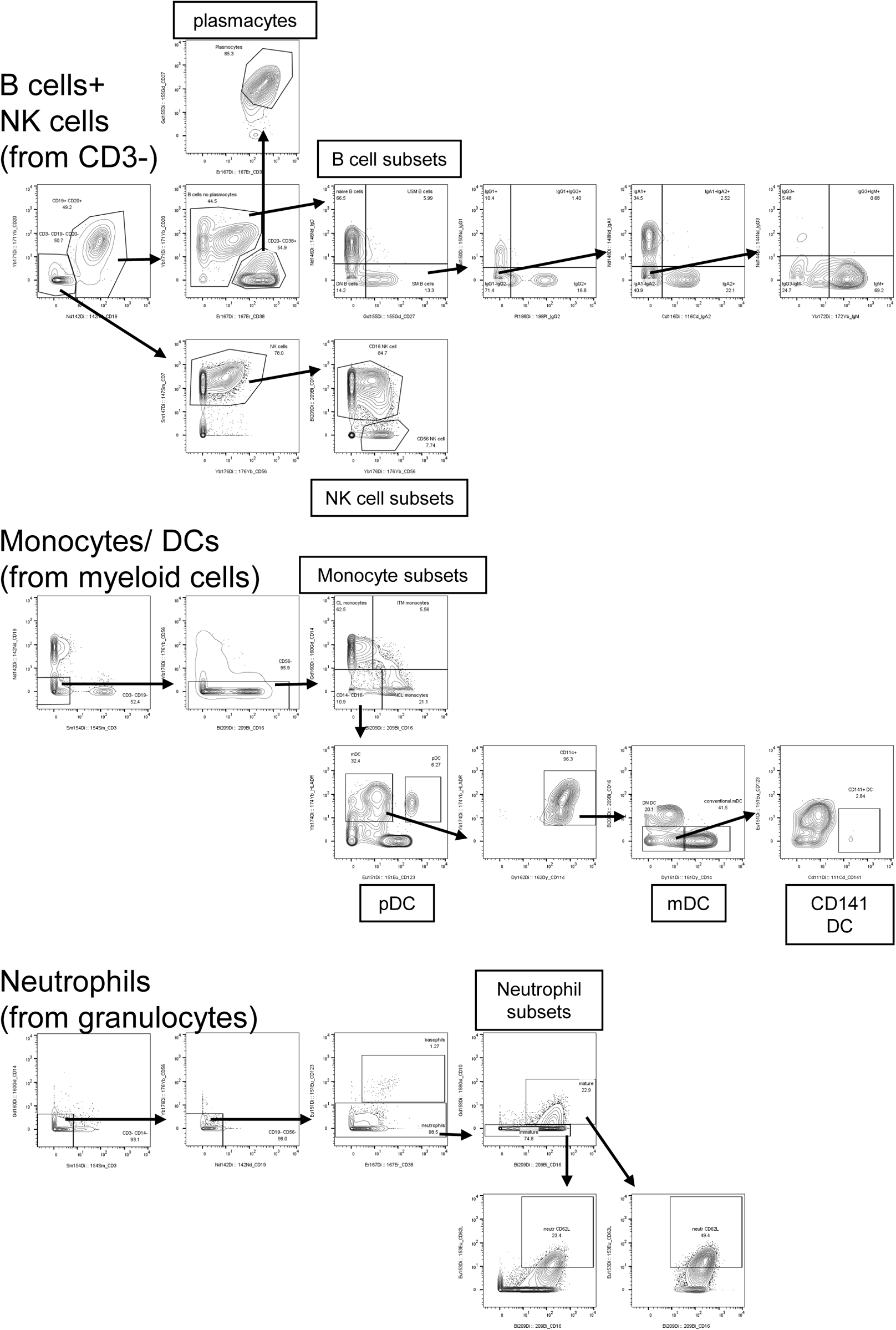
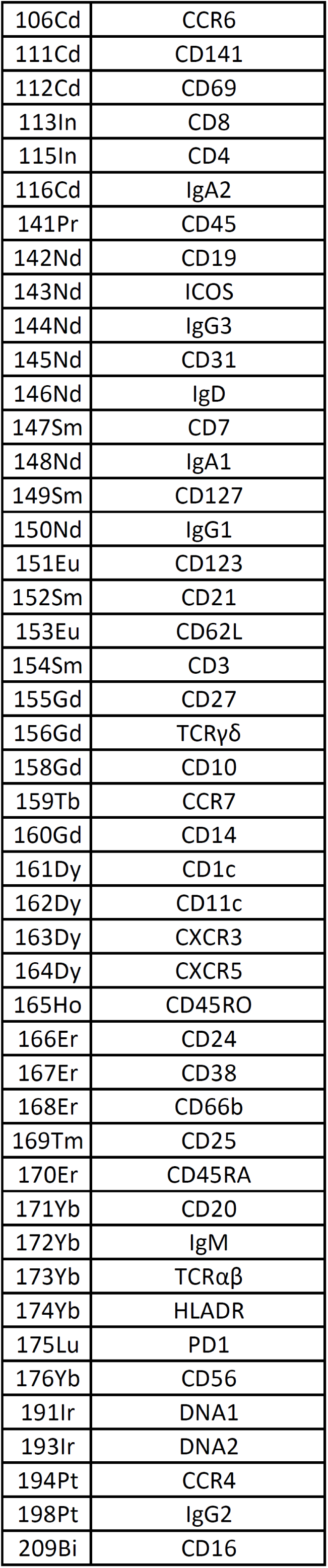
Gating strategy for mass cytometry. Complete gating strategy identify the main population with the list of antibodies and coupled antibodies.

**Supplementary Table 1:**
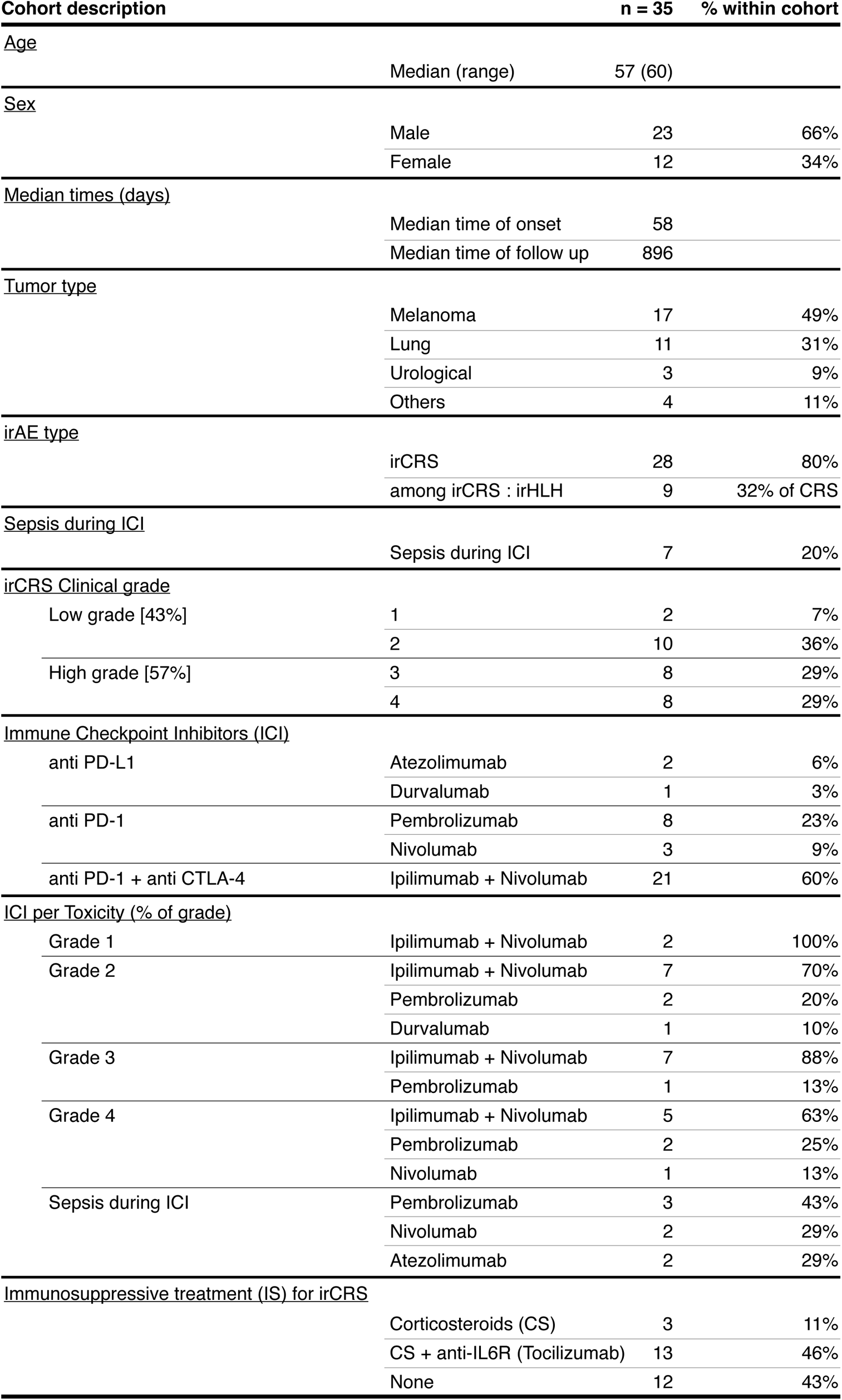
Patients, treatments, and clinical characteristics. The principal clinical, demographic, and therapeutic data for the entire cohort (n=35) are presented in detail in this table.

**Supplementary Table 2:** clinical characteristics of irCRS patients receiving Tocilizumab treatment (n=12).

| Patients Receiving Tocilizumab |  | n (N=12) | % |
| --- | --- | --- | --- |
| <u>Sex</u> |  |  |  |
|  | Male | 6 | 50 |
|  | Female | 6 | 50 |
| <u>Age</u> |  |  |  |
|  | Median (Mean) | 46 (48) |  |
| <u>Mortality related to irCRS</u> |  |  |  |
|  | 7 days | 0 | 0.00 |
|  | 30 days | 0 | 0.00 |
| <u>Clinical Grading</u> |  |  |  |
|  | irCRS Grade 3 | 5 | 41.67 |
|  | irCRS Grade 4 | 7 | 58.33 |
| <u>irHLH</u> |  |  |  |
|  | irHLH within Grade 3 | 1 | 20.00 |
|  | irHLH within Grade 4 | 7 | 100.00 |
| <u>Severity factors</u> |  |  |  |
|  | Acute respiratory failure | 5 | 41.67 |
|  | Acute Kidney Injury | 7 | 58.33 |
|  | Acute Liver failure | 2 | 16.67 |
|  | Admission in Intensive or Intermediate Care | 5 | 41.67 |
|  | DIC | 6 | 50.00 |
|  | Fluid resuscitation | 5 | 41.67 |
|  | Vasopressors | 2 | 16.67 |
|  | High dose of steroids (HDS) | 12 | 100.00 |
| <u>Mortality related to irHLH</u> |  |  |  |
|  | 7 days | 0 | 0.00 |
|  | 30 days | 0 | 0.00 |
| <u>irHLH response</u> |  |  |  |
|  | Complete Resolution | 12 | 100.00 |
| <u>Oncological response 3 months after TCZ</u> |  |  |  |
|  | Complete Response (CR) | 3 | 25.00 |
|  | Stable Disease (SD) | 2 | 16.67 |
|  | Partial Response (PR) | 0 | 0.00 |
|  | Progressive Disease (PD) | 6 | 50.00 |
|  | Not Assessed | 1 | 8.33 |
| <u>Oncological response 6 months after TCZ</u> |  |  |  |
|  | Complete Response (CR) | 4 | 33.33 |
|  | Stable Disease (SD) | 2 | 16.67 |
|  | Partial Response (PR) | 0 | 0.00 |
|  | Progressive Disease (PD) | 3 | 25.00 |
|  | Not Assessed | 3 | 25.00 |

**Supplementary Table 3: Global biomarker table.** Summary table with for irCRS G1-G4, Bacterial sepsis, Viral sepsis, Immunosuppressed patients: N, Mean, Min, Max, Percentiles (25th,50th,75th) and SD. Lower limit of detection is included in the table for all biomarkers.

**Supplementary Table 4:**
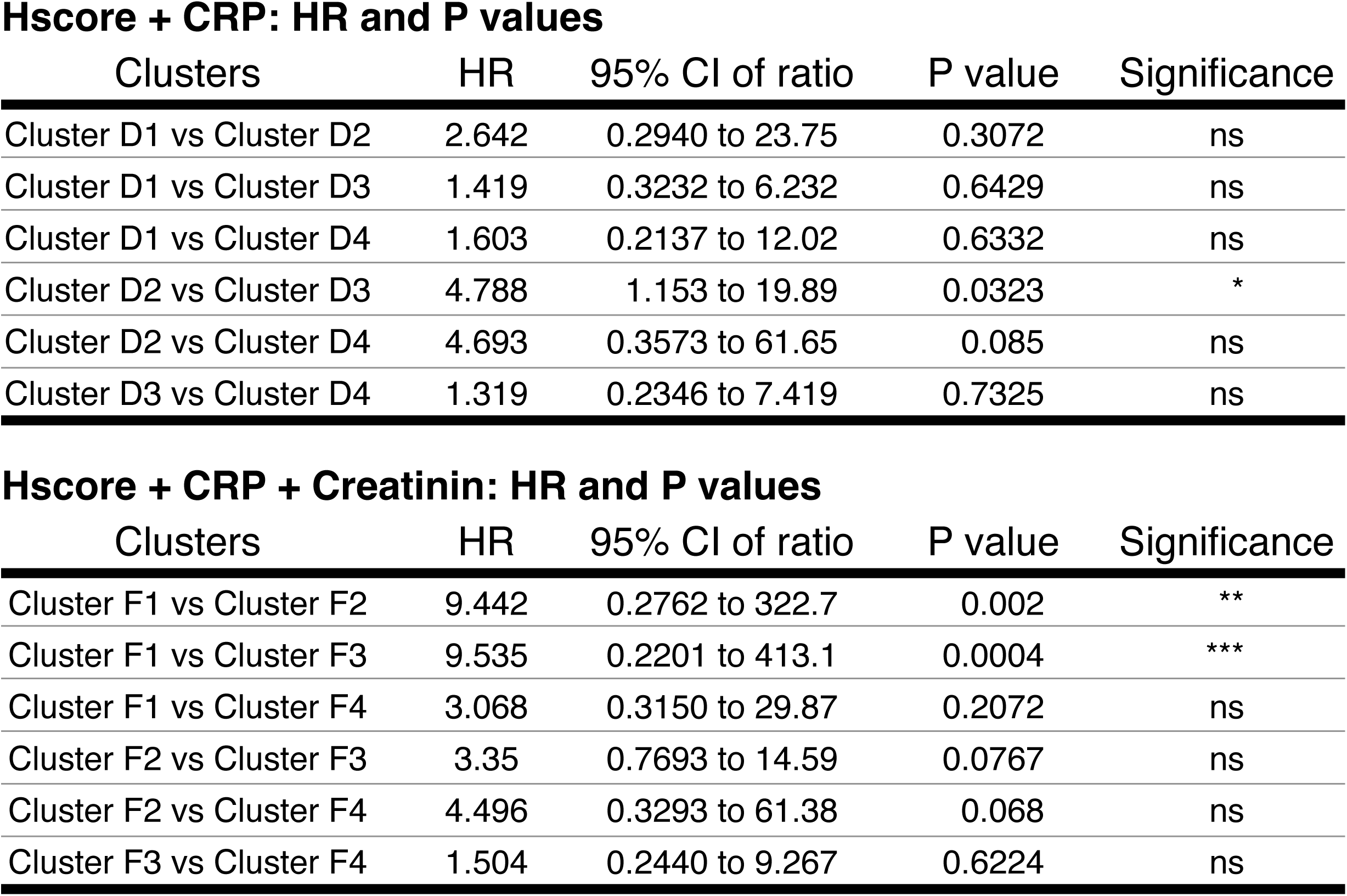
HR and P values associated with Figure 2 panels D and F. logrank test for comparing clusters in Figure 2 panels D and F.

Hscore + CRP: HR and P values
| Clusters | HR | 95% CI of ratio | P value | Significance |
| --- | --- | --- | --- | --- |
| Cluster D1 vs Cluster D2 | 2.642 | 0.2940 to 23.75 | 0.3072 | ns |
| Cluster D1 vs Cluster D3 | 1.419 | 0.3232 to 6.232 | 0.6429 | ns |
| Cluster D1 vs Cluster D4 | 1.603 | 0.2137 to 12.02 | 0.6332 | ns |
| Cluster D2 vs Cluster D3 | 4.788 | 1.153 to 19.89 | 0.0323 | * |
| Cluster D2 vs Cluster D4 | 4.693 | 0.3573 to 61.65 | 0.085 | ns |
| Cluster D3 vs Cluster D4 | 1.319 | 0.2346 to 7.419 | 0.7325 | ns |

Hscore + CRP + Creatinin: HR and P values
| Clusters | HR | 95% CI of ratio | P value | Significance |
| --- | --- | --- | --- | --- |
| Cluster F1 vs Cluster F2 | 9.442 | 0.2762 to 322.7 | 0.002 | ** |
| Cluster F1 vs Cluster F3 | 9.535 | 0.2201 to 413.1 | 0.0004 | *** |
| Cluster F1 vs Cluster F4 | 3.068 | 0.3150 to 29.87 | 0.2072 | ns |
| Cluster F2 vs Cluster F3 | 3.35 | 0.7693 to 14.59 | 0.0767 | ns |
| Cluster F2 vs Cluster F4 | 4.496 | 0.3293 to 61.38 | 0.068 | ns |
| Cluster F3 vs Cluster F4 | 1.504 | 0.2440 to 9.267 | 0.6224 | ns |

**Supplementary Table 5:** Clinical characteristics of irCRS patients within clusters identified in Figure 3.

|  | Cluster 1<br>(n=7) | Cluster 2<br>(n=18) | Cluster 3<br>(n=3) |
| --- | --- | --- | --- |
| <u>Grading irCRS/irHLH</u> |  |  |  |
| 1 | 0 | 2 | 0 |
| 2 | 0 | 9 | 0 |
| 3 | 4 | 4 | 0 |
| 4 | 3 | 2 | 3 |
| ... irHLH Patients | 4 | 2 | 3 |
| <u>Cancer type</u> |  |  |  |
| Breast | 0 | 1 | 0 |
| Digestive | 0 | 2 | 0 |
| Lung | 0 | 7 | 2 |
| Melanoma | 7 | 8 | 0 |
| Genitourinary | 0 | 0 | 1 |
| <u>Treatment</u> |  |  |  |
| TCZ + GC | 6 | 5 | 2 |
| GC | 1 | 1 | 1 |
| No Immunosuppressors | 0 | 11 | 0 |

**Supplementary Table 6:** ROC curve analysis of predictors of irHLH versus irCRS-G3/Sepsis patients. ROC, AUC and p values, cutoff, Sensitivity, Specificity, Accuracy, Positive predictive value and Negative predictive value.

| irHLH vs irCRS-G3 & Sepsis Predictors |  |  |  | Sensitivity |  | Specificity |  | Accuracy | PPV | NPV |
| --- | --- | --- | --- | --- | --- | --- | --- | --- | --- | --- |
| Biomarker | ROC AUC | ROC P value | Cutoff | % | 95% CI | % | 95% CI |  |  |  |
| Ferritin | 1 | 0.0009 | < 23221 | 100 | 74.12% to 100.0% | 100 | 60.97% to 100.0% | 100.00 | 100.00 | 100.00 |
| CCL4 | 0.9848 | 0.0013 | < 164.0 | 90.91 | 62.26% to 99.53% | 100 | 60.97% to 100.0% | 94.12 | 85.71 | 100.00 |
| HGF | 0.9848 | 0.0013 | < 592.5 | 90.91 | 62.26% to 99.53% | 100 | 60.97% to 100.0% | 94.12 | 85.71 | 100.00 |
| IL-1RA | 0.9697 | 0.0018 | < 12251 | 100 | 74.12% to 100.0% | 83.33 | 43.65% to 99.15% | 94.12 | 100.00 | 91.67 |
| IL-10 | 0.9697 | 0.0018 | < 25.00 | 100 | 74.12% to 100.0% | 83.33 | 43.65% to 99.15% | 94.12 | 100.00 | 91.67 |
| IL-18 | 0.9697 | 0.0018 | < 391.0 | 100 | 74.12% to 100.0% | 83.33 | 43.65% to 99.15% | 94.12 | 100.00 | 91.67 |
| SCF | 0.947 | 0.003 | < 21.00 | 90.91 | 62.26% to 99.53% | 83.33 | 43.65% to 99.15% | 88.24 | 83.33 | 90.91 |
| aPTT | 0.93 | 0.0085 | < 43.85 | 100 | 72.25% to 100.0% | 80 | 37.55% to 98.97% | 93.33 | 100.00 | 90.91 |
| CCL2 | 0.9242 | 0.0049 | < 211.5 | 90.91 | 62.26% to 99.53% | 83.33 | 43.65% to 99.15% | 88.24 | 83.33 | 90.91 |
| D-Dimers | 0.92 | 0.0101 | < 12708 | 80 | 49.02% to 96.45% | 100 | 56.55% to 100.0% | 86.67 | 71.43 | 100.00 |
| Fibrinogen | 0.9091 | 0.0067 | > 1.450 | 100 | 74.12% to 100.0% | 83.33 | 43.65% to 99.15% | 94.12 | 100.00 | 91.67 |
| CXCL9 | 0.9091 | 0.0067 | < 357.5 | 100 | 74.12% to 100.0% | 83.33 | 43.65% to 99.15% | 94.12 | 100.00 | 91.67 |
| Prothrombin Ratio | 0.8939 | 0.009 | > 65.00 | 100 | 74.12% to 100.0% | 66.67 | 30.00% to 94.08% | 88.24 | 100.00 | 84.62 |
| CXCL10 | 0.8939 | 0.009 | < 196.5 | 90.91 | 62.26% to 99.53% | 83.33 | 43.65% to 99.15% | 88.24 | 83.33 | 90.91 |
| CCL3 | 0.8712 | 0.0138 | < 8.500 | 90.91 | 62.26% to 99.53% | 83.33 | 43.65% to 99.15% | 88.24 | 83.33 | 90.91 |
| IFN-γ | 0.8636 | 0.0159 | < 16.50 | 63.64 | 35.38% to 84.83% | 100 | 60.97% to 100.0% | 76.47 | 60.00 | 100.00 |
| Platelets | 0.8485 | 0.0208 | > 79.00 | 81.82 | 52.30% to 96.77% | 83.33 | 43.65% to 99.15% | 82.35 | 71.43 | 90.00 |
| CXCL13 | 0.8333 | 0.027 | < 440.0 | 90.91 | 62.26% to 99.53% | 83.33 | 43.65% to 99.15% | 88.24 | 83.33 | 90.91 |
| CCL11 | 0.803 | 0.0444 | < 25.50 | 63.64 | 35.38% to 84.83% | 100 | 60.97% to 100.0% | 76.47 | 60.00 | 100.00 |

**Supplementary File 1:** P values for comparison of mass cytometry data comparing either Low grade or high-grade patients with baseline. Statistical significance tested with Mann-Whitney test.

**Supplementary File 2:** P values for comparison of mass cytometry data comparing either high grade or sepsis patients with baseline. Statistical significance tested with Mann-Whitney test.

## Material and Methods

### Sample collection and ethic approval

All patients either signed informed consent offered to all patients to allow research use of their data in a coded fashion (called “*consentement general*”) or did not express opposition and were included thanks to article 34 of the Swiss federal law relative to human research. Research protocol was approved by the cantonal ethical committee “*Commission cantonal d’éthique de la recherche sur l’être humain (CER-VD)”.* All samples were harvested during the normal clinical practice, and no specific intervention on patients were performed regarding this study.

### Immune profiling of circulating blood immune cells population by mass cytometry

Patient blood was stained as previously described (*37*). Briefly, cells were incubated for 30 min at room temperature (RT) with a 50μL antibody cocktail of metal-conjugated antibodies. Cells were then washed and fixed with 2.4% PFA for 10 min at RT, lysed for 15 min at RT using Bulklysis solution (Cytognos), and incubated 30 min at RT with metal-conjugated antibodies. Complete list of metal-conjugated antibodies and gating strategy can be found as Supplementary Figure 9. Cells were washed and total cells were identified by DNA intercalation (1μM Cell-ID Intercalator, Standard BioTools) in 1.6% PFA at 4 °C overnight. Labeled samples were acquired using the HELIOS CyTOF system (Standard BioTools) and FCS files were normalized to EQ Four Element Calibration Beads using the CyTOF software.

### Immune profiling of serum biomarkers

As previously described (*38*) serum concentrations of cytokines, soluble CD25, chemokines, and growth factors were determined by Luminex ProcartaPlex immunoassays for each marker. List of biomarkers, lower limit of detection and distribution of values within our cohort can be found in **Supplementary Table 3**. Samples values below or equal to the LLOD were replaced by the LLOD.

### Statistical analysis

All statistical analysis in figures comparing cytokines were performed using GraphPad Prism 10.1.2. Most graphs were also done using GraphPad Prism, in exception of correlation figures, and machine learning figures that were done using R. Mass cytometry analysis was performed using FlowJo. 1-d k-mean clusters were down using the ckmeans.1.dp R packages (Song and Zhong, 2020) and optimal number of cluster evaluated using silhouette method. sPLS-DA was performed using the “mixOmics” package and random forest using the R package “randomForest”.). Statistical analysis between groups have been done using either student T test, Mann-Whitney U test or 1-way ANOVA as indicated in figures. All p values below 0.05 were defined as significant.

### Custom code availability

All custom code used for analysis was written using R or Matlab R2023b. Custom code example is available on the LCIT github repository (https://github.com/LCIT-CHUV/ImmunoTox).

### Data Availability

The datasets supporting the results of this study are not publicly available. Requests for access to the dataset will be granted upon reasonable request to the principal investigator. Study data will be managed, stored, shared, and archived according to CHUV standard operating procedures to ensure the continued quality, integrity, and utility of the data.

## Discussion

Our comprehensive study examines the immune dynamics of immune-related cytokine release syndrome (irCRS) and reveals distinct patterns across CRS grades. We found that CXCL9, CXCL10 and IFN-γ levels increase with CRS severity, suggesting their role as early markers of irCRS onset and progression. This finding underscores the importance of monitoring these markers for early intervention, potentially reducing the severity of irCRS and improving outcomes. When comparing irHLH patients to those with grade 3 irCRS, we found significant elevations in proinflammatory and antitumor cytokines (including IFN-γ, IL-6, IL-18, CXCL9, CXCL10, CXCL13, CCL2, CCL3, CCL4, HGF, SCF, and PDGF-B), along with increases in anti-inflammatory cytokines (IL1-RA, IL-10). This cytokine profile suggests a complex inflammatory continuum at the intersection of irCRS severity, irCARS, and oncologic response, highlighting the nuanced immune responses induced by ICI therapy. Our study reveals the simultaneous emergence of irCARS and irCRS during ICI treatment, characterized by increased anti-inflammatory cytokines, particularly IL1-RA and IL-10. This observation highlights a dynamic balance between pro-inflammatory and anti-inflammatory responses that intensifies as CRS progresses. Specifically, we observed a significant increase in IL-10 at grade 4, suggesting its role in amplifying the irCARS response under intense inflammatory condition. We also documented significant correlations between ferritin, d-dimer, and Hb and IL-6, illustrating the complex relationship between CRS severity, anemia, and coagulopathy. Furthermore, an increase in IL-1 family cytokines (IL-1β, IL-18, IL-1RA) during ICI therapy suggests inflammasome activation, particularly NLRP3, indicating its substantial influence on the CRS and CARS continuum. NLRP3 inflammasome was reported for its role in processing and releasing pro-inflammatory cytokines, and its significant influence on the inflammatory continuum of CRS and CARS (*26*).

The dominant Th1 response associated with ICI, characterized by significant IFN-γ production without substantial involvement of IFN-α or Th17 pathways, suggests a specific immune activation pathway critical for the therapeutic efficacy of ICI. This is further supported by the elevated levels of IL-10, IL1-RA, CXCL9 and CXCL10 in cluster 1 with improved OS highlighting these markers as potential biomarkers of enhanced anti-tumor activity in ICI therapy. Specifically, these inflammatory chemokines bind to the CXCR3 receptor and specifically target activated T lymphocytes and natural killer (NK) cells (*39–41*). A pronounced increase in HGF levels and in IL-10/HGF correlation in patients with severe irAEs (grade 3 and 4) suggests its role as a biomarker for irCARS and for clinical severity. The correlation between HGF levels and irCARS amplification highlights HGF’s potential in moderating inflammatory responses, possibly aiding in tissue repair and inflammatory damage mitigation (*42*). This observation opens avenues for exploring HGF’s role in both irCRS and irCARS, particularly in the context of ICI therapy.

Our findings underscore the importance of multi-biomarker strategies in refining survival predictions and tailoring therapeutic interventions for irCRS. Notably, we observed contrasting immune patterns between irHLH clusters. Cluster 3, associated with high acute mortality due to irHLH rather than cancer progression, showed elevated inflammatory cytokines (IL-6, IL-8, CXCL8, CXCL13, IL1-RA, HGF, SCF) compared to clusters 1 and 2, which showed better OS and higher levels of CXCL9, CXCL10, IFN-γ, and IL-10. This indicates a broader inflammatory response in irHLH and suggests that robust irCARS activation could counterbalance intense inflammation. The distinction between the cytokine profiles in these clusters underscores the complexity of irAEs in irHLH and suggests that more regulated inflammatory responses in clusters 1 and 2 may lead to improved outcomes. Elevated pro-inflammatory cytokines in cluster 3 suggest a cytokine milieu contributing to the severity of irHLH that is distinct from clusters focused on oncologic responses. This analysis highlights the need for tailored therapeutic strategies based on cluster-specific cytokine patterns and underscores the critical nature of managing the extensive organ involvement and systemic impact seen in severe irHLH cases. Our comprehensive analysis highlights the importance of personalized therapeutic approaches to mitigate adverse outcomes in severe immune-mediated diseases, which is essential to optimize ICI therapy management, especially in patients with severe irAEs such as irHLH.

The rationale of our study is based on the observed dichotomy between the Lee criteria for CRS grading and the HScore in assessing CRS severity and HLH characteristics. We found significant discrepancies between these criteria, noting cases of high-grade irCRS by Lee criteria with unexpectedly low HScore, and vice versa. This highlights the complexity of categorizing irCRS and irHLH and suggests a continuum between them, ranging from purely biological to mixed clinical manifestations. Our analysis suggests that the HScore alone may inaccurately represent irCRS G3 and irHLH, leading to potential mismanagement based on its sensitivity of approximately 71%. These findings underscore that a high HScore doesn’t always warrant aggressive interventions, such as high-dose steroids, typically prompted by high Lee grades. Cases in which a lower HScore is associated with severe CRS challenge the stand-alone utility of the HScore to guide immunosuppressive therapy. Our goal was to identify markers that are more predictive of clinical severity and appropriate initiation of therapy. This approach acknowledges the insights of the HScore, but argues against its sole use for critical decisions, advocating a holistic interpretation that considers the broader clinical context and patient phenotypes. Integrating the Lee criteria and HScore with new biomarkers may lead to more personalized, effective treatment strategies for irCRS and irHLH, using both scores and a comprehensive assessment of clinical and inflammatory markers to improve diagnostic accuracy and interventional guidance.

In evaluating 45 biomarkers, our study identified HGF and ferritin as particularly predictive of irHLH versus irCRS G3, with 100% sensitivity and specificity. Biomarkers associated with severe irCRS (grade 4) showed strong discriminatory power such as SCF, IL-6, IL-1RA, CCL2, CXCL9, CXCL13, ASAT, fibrinogen, D-dimer, and prothrombin ratio, while others such as lymphocyte count and hemoglobin were less effective. In particular, CXCL9 stood out for its dual role in distinguishing irHLH from irCRS and predicting the need for treatment intensification with TCZ, underscores the potential for tailored therapeutic strategies. Similarly, HGF, SCF, IL-1RA and others proved valuable in predicting clinical needs such as fluid resuscitation, improving patient management and guiding novel treatments. Similarly, HGF, SCF, IL-1RA, CCL2, IL-18, ferritin, prothrombin ratio, and D-dimer showed dual performance in predicting hemodynamic instability requiring fluid resuscitation and in distinguishing irHLH from irCRS. The potential of these biomarkers to predict critical clinical outcomes, such as hemodynamic instability further underscores their utility in clinical practice.

Recent studies are consistent with our findings and showed distinct cytokine profiles in irHLH compared to sepsis, with increased levels of CXCL9, CXCL10, and CXCL11 in HLH versus sepsis, while IL-6 was relatively higher in SIRS/sepsis compared to HLH, and IL-10 showed no significant difference(*43*). In addition, we noted a distinct cytokine signature in irHLH, significantly differing from sepsis with elevated IL-10, IL-18, IFN-γ, CCL2, CCL3, HGF, SCF, and IL1-RA, along with ferritin and d-dimer. IL-2R did not distinguish between irHLH and sepsis. Dual inflammatory and biological signatures differentiated the irHLH, sepsis, and grade 3 CRS groups. IL-10, HGH, IL-18, IL1-RA, and SCF were key in irHLH, while leukocytes, IL-7, EGF, PDGF-BB, and GM-CSF were prominent in sepsis. Our data suggest that ferritin, IL-10, EGF and total leukocytes can discriminate between irHLH, irCRS-3 and sepsis. These biomarkers achieved 100% accuracy in our data, highlighting the potential of these biomarkers in clinical practice for early diagnosis, patient stratification and tailored therapies.

Recent studies have shown that T cell activation profiles can discriminate between hyperinflammatory disorders such as HLH and sepsis, with >7% CD38^high^/HLA-DR+ among CD8+ T cells being a key discriminator (*44*). This was extended to CD38^high^/HLA-DR+CD8+ T cells expressing CD4 (CD4^dim^CD8+ T cells), which correlated with HLH severity and levels of CXCL9 and IL-18 (*45*). Our research explores these immunologic profiles and reveals increased CD38+/HLA-DR+ cell frequencies across multiple T cell subtypes, indicating a specific and robust immune memory response in irCRS, possibly due to prior antigen exposure or the nature of the immune stimulus. This distinct T-cell profile may differentiate irCRS from sepsis and measure the severity of the immune response in patients undergoing ICI therapy.

In addition, we found that neutrophils and monocytes in irCRS patients exhibited more CD38, CD11c, and CD62L upregulation than in sepsis, suggesting a different activation profile that could help differentiate irCRS from sepsis. This indicates a different inflammatory and immunoregulatory environment in irCRS, highlighting the potentially unique role of innate immune cells in its pathogenesis. The specific activation of monocytes, which are critical for antigen presentation and cytokine production, highlights their importance in the severity of irCRS and as possible targets for therapeutic intervention or disease monitoring markers.

The significant expansion of our patient cohort from three cases in our previous report (*15*) to the inclusion of twelve patients in this study underscores the growing body of evidence supporting the efficacy of TCZ in the treatment of CS-refractory high-grade irCRS. This larger patient cohort not only reaffirms but also strengthens our confidence in the important therapeutic potential of TCZ.

This study has several limitations, including a small cohort size and a limited number of paired samples, which may affect the generalizability of the findings. As a single-center study, it may be subject to selection bias and heterogeneity due to the patient population, differences in disease stage, prior treatments, and comorbidities. This limitation underscores the importance of building larger cohorts to address these confounding factors more accurately. These factors could influence the results of the study. In addition, the lack of validation in independent cohorts limits the reproducibility and robustness of our conclusions. Longitudinal data collection from baseline and during ICI treatment could improve data quality and reliability.

Despite these limitations, our findings advocate a paradigm shift in the diagnostic approach to immune-related adverse events and propose a multi-biomarker strategy that not only improves diagnostic accuracy but also aids in the prognosis and management of irCRS and irHLH and their distinction from sepsis. This approach promises a significant step toward personalized medicine. It will enable clinicians to predict treatment response and tailor interventions with unprecedented accuracy. Ultimately, this research provides the foundation for a new era in managing irCRS and irHLH characterized by increased diagnostic accuracy, improved patient outcomes, and personalized therapeutic interventions.

## Acknowledgements

This work was supported by the strategic plan of the CHUV. We would like to express our gratitude to all the patients who generously contributed their time and samples for this project. We would like to thank Pr Gérard Waeber and Pr Peter Vollenweider for their help in the clinical care of our patients. We would like to thank Federica Martina for her initial version of the code used in the article. Visual abstract was created with BioRender.com.

## Author contributions

M.O. and D.D. had full access to all data in the study and take responsibility for its integrity and accuracy. MO conceived, designed the study and drafted the manuscript. D.D., G.P., and M.O. analyzed and interpreted the data. D.D., A.S., and M.O. collected the data. S.L., N.M., H.B., R.D., K.A., N.F., J.D., G.S., L.M., V.M., D.B., A.S., C.S., K.S. and S.P. participated in the clinical treatments. V.J., R.B. and AN analyzed the CyTOF data. D.D. and M.O. prepared the figures. J.T. and Y.W. participated in scientific discussion. The manuscript was reviewed and approved by all authors before submission.

## Competing interests

MO received honoraria and speaker fees from Moderna, Roche and BMS.

## Notes

### Funding Statement

The study was funded by CHUV institutional funding

### Author Declarations

Ethics committee of the Canton of Vaud (Commission cantonal d'ethique de la recherche sur l'etre humain (CER-VD)) gave ethical approval for this work.

